# Deep Learning in Dentistry: A Systematic Review from an AI Researcher Viewpoint

**DOI:** 10.1101/2025.10.01.25337082

**Authors:** Zhi Qin Tan, Marina Guimaraes Roscoe, Owen Addison, Yunpeng Li

**Affiliations:** Centre for Oral, Clinical & Translational Sciences, Faculty of Dentistry, Oral & Craniofacial Sciences, King’s College London, London, UK

**Keywords:** Artificial Intelligence, Caries detection/diagnosis/prevention, Deep Learning/Machine Learning, Dental anatomy, Dental informatics/bioinformatics, Periodontal disease(s)/periodontitis

## Abstract

**Background:** Deep learning has achieved rapid development in recent years and has been applied to various fields in dentistry. While cross-disciplinary research between artificial intelligence and dentistry is growing exponentially, most studies rely on off-the-shelf machine learning models, with only a small portion introducing technological novelty. Furthermore, tasks such as dental disease diagnosis are inherently complex, with high intra- and interobserver variability where dentists often interpret radiographs differently and offer varying subsequent treatments. However, many studies overlooked this variability, assuming no data and model uncertainty in dental tasks. Additionally, many evaluated their methods using private and small-scale datasets, making fair comparisons of their outcome metrics challenging and introducing significant predictive bias in artificial intelligence models. The goal of the current study was to examine and critically assess recent novel advances in artificial intelligence in dentistry across a wide range of dental applications.

**Methods:** We begin by presenting foundational concepts in artificial intelligence and adopt a unique approach by focusing on the novelty of deep learning methods. Following that, we conducted a systematic review by searching online databases (PubMed, IEEE Xplore, arXiv, and Google Scholar) for publications related to artificial intelligence, machine learning, and deep learning applications in dentistry.

**Results:** A total of 91 articles met the inclusion criteria, and we presented a comprehensive analysis of the studies. Moreover, we discuss the limitations of recent studies on artificial intelligence in dentistry and identify key research opportunities for progress and innovation. These include integrating dental domain knowledge, quantifying uncertainty, leveraging large models and multiple sources of datasets, developing efficient deep learning pipelines, and conducting thorough evaluations in both simulated and real-world experimental settings.

**Conclusion:** Recent advancements in deep learning demonstrate great potential in dentistry applications. However, future research to address the limitations in recent studies is needed to fully realize its potential for enhancing dental professionals to utilize AI effectively and improve clinical and patient outcome in dentistry.

## 1 Introduction

Artificial intelligence (AI) involves developing computer programs that solve problems that traditionally require human intelligence. Applications range from basic expert systems that mimic human decision-making through pre-defined rules to more advanced tools such as ChatGPT which can engage in contextual conversations. In recent years, AI has achieved significant advances in numerous machine learning tasks and has been increasingly adopted across various fields including healthcare (1).

In dentistry, AI has rapidly progressed to improve diagnostic accuracy, prognosis prediction, and process efficiency. In most cases, AI serves as an assistive tool, enabling clinicians to allocate more time to high-value tasks surrounding patient interaction and treatment provision. Compared with conventional machine learning techniques, such as decision trees and support vector machines, the use of deep learning techniques among the dentistry community has increased significantly. Most reviews on AI applications in dentistry, written from the perspective of clinical researchers (2–7) offer valuable insights but face methodological challenges including unclear definitions of fundamental deep learning concepts. Furthermore, recent articles lack the discussion of data and model uncertainty, reported results with private small datasets, and have yet to explore large AI models, a trend that deviates from the norm in computer science research. This review adopts a unique perspective from computer scientists, aiming to clarify several key AI terms and critically review recent advances in dental AI. Definitions are provided in Table 1, followed by an exploration of deep learning applications in dentistry.

**Table 1:** Glossary of technical terms used in this paper.

| Term | Definition |
| --- | --- |
| Computer vision | The field of study in computer science involving processing, analyzing and understanding of digital images/videos to derive information. |
| Natural language processing | The field of study in computer science that uses artificial intelligence to enable computers to understand and manipulate human texts and speeches. |
| Decision tree | A tree-like model that sequentially partitions the input domain into possible outcomes using if-else rules in its branches. |
| Support vector machine | A supervised machine learning algorithm that classifies data by determining the optimal line/hyperplane that separates all classes. |
| Deep learning | The subfield of study in machine learning that utilizes artificial neural networks (ANN) to process data and make predictions, mimicking the human brain. |
| Representation learning | A process in machine learning that allows algorithms to extract meaningful high-level features from raw data. |
| High-level feature | Attributes extracted from data that are more meaningful/representative than low-level features, e.g. multiplication of two input features. |
| Nodes of ANN | Simple processing units that receive inputs from nodes in the previous layer and output the results to the next layer of nodes. |
| Weights of ANN | Learnable parameters that represent the strength/importance of the connection of each pair of nodes between two consecutive layers. |
| Forward pass | The calculation process of ANN by passing the data from the input nodes until the output nodes to produce predictions. |
| Activation function | A function that transforms the output of nodes to introduce non-linearity into the ANN to enable it to learn complex patterns in the data. |
| Loss function | A function that quantifies the model’s error and is used during the optimization process where the weight parameters are learnt by minimizing it. |
| Gradient descent | An optimization algorithm that iteratively finds the minimum of a given function by moving in the opposite direction of its gradient. |
| Convolutional neural network (CNN) | A variant of ANN designed for images by utilizing convolution layers (2-dimension matrix of nodes) instead of layers of single dimension nodes. |
| Recurrent neural network (RNN) | A variant of ANN designed for sequential data (e.g. texts, audio) by incorporating an internal state to retain information of past inputs. |
| Transformer models | A variant of ANN incorporating a self-attention mechanism, eliminating the need of a recurrent process of RNN for sequential data. |
| c-class classification | Classification task to classify data into c classes. |
| SoftMax function | A function to rescale an n-dimensional input so that they emit a valid categorical probability distribution, i.e. they sum to one. |
| Cross-entropy loss | A loss function for classification tasks that measures the difference between the predicted class probability and the true distribution of the labels. |
| Object predictions | A set of predictions of object instances, represented by the predicted class labels, bounding box locations and confidence scores. |
| Precision-recall curve | A plot of precision as a function of recall for all possible confidence score thresholds, illustrating the tradeoff between precision and recall. |
| Mask R-CNN | An extension of the Faster R-CNN model that can perform object detection and instance segmentation simultaneously. |
| Generative adversarial network (GAN) | A deep learning framework for estimating generative models in which two neural networks compete against each other in an adversarial manner. |
| Generator neural network | The generative model in the GAN framework that creates plausible data from random input by incorporating feedback from the discriminator model. |
| Discriminator neural network | The classification model in the GAN framework that distinguishes real data from the data generated by the generator model. |
| Split bias | A dataset bias because of splitting the dataset into training and testing sets causing different data characteristics of the split sets. |
| Level set algorithm | A classical image segmentation technique that is based on the differences of neighboring pixels to find object boundaries progressively. |
| Attention module | A mechanism introduced to improve performance of transformers, allowing them to determine the relative importance between each component of the sequential data. |
| Multi-scale spatial pyramid pooling | A CNN building block that allows models to capture spatial features at multiple image scales/sizes. |
| Squeeze and excitation module | A CNN architectural unit that allows model to learn channel-wise importance weights of inputs. |
| Cascaded CNN | A type of modeling approach that implements multiple CNN that typically operates at multiple resolutions to iteratively improve model predictions. |
| Capsule network | A variant of ANN that attempts to better model the hierarchical relationships by using sets of nodes (capsules) to represent object properties in an image. |
| Multi-head attention mechanism | An approach of using several attention modules in parallel to allow for attending to different parts of an input sequence differently. |
| Full-scale axial attention module | A variant of the attention module that allows capturing of the full dependencies of high dimensional data (e.g. images). |
| Teacher-student model | A knowledge distillation technique to transfer learnt knowledge from a larger teacher model to a smaller and more efficient student model. |
| Parallel convolutional layers | A modeling approach in CNN that uses multiple parallel convolutional branches to extract features at multiple locations and sizes simultaneously. |
| Local and global branches | Dual-branch modeling approach in CNN to capture both global (whole image) and local (region of interest) features of an image. |
| Relational reasoning network | A network designed to solve problems related to relational reasoning by learning rich relationship among different entities in the input. |
| Foundation model | A large machine learning model (typically a deep learning model) trained on vast data that can be adapted to various downstream use cases and tasks. |

### 1.1 Machine Learning

Machine learning is a subset of AI that develops and studies statistical algorithms to enable computer programs to perform tasks without explicit instructions from humans. It can be broadly categorized into three categories:

1. Supervised learning: The algorithm learns to map inputs to outputs using labeled examples.
2. Unsupervised learning: The algorithm finds patterns in input data without labeled outputs.
3. Reinforcement learning: The algorithm learns by interacting with an environment and maximizing rewards based on feedback.

### 1.2 Deep Learning

Deep learning is a type of machine learning method based on artificial neural networks (ANNs) with representation learning. Deep learning automates feature extraction using multiple layers of nodes and weights that progressively identify high-level features for complex tasks (Figure 1). Generally, the learning of the weight parameters is performed by first presenting the inputs to the ANN and obtaining the model outputs. In the forward pass, input data moves through intermediate layers, applying non-linear activation functions to produce the final output. Using gradient descent, weight parameters are updated to minimize a loss function, a process called back-propagation. These steps are repeated until the model reaches satisfactory accuracy. Variants of artificial neural networks (ANNs) like convolutional neural networks (CNNs), recurrent neural networks (RNNs), and transformer models have been developed for various applications.

**Figure 1:**
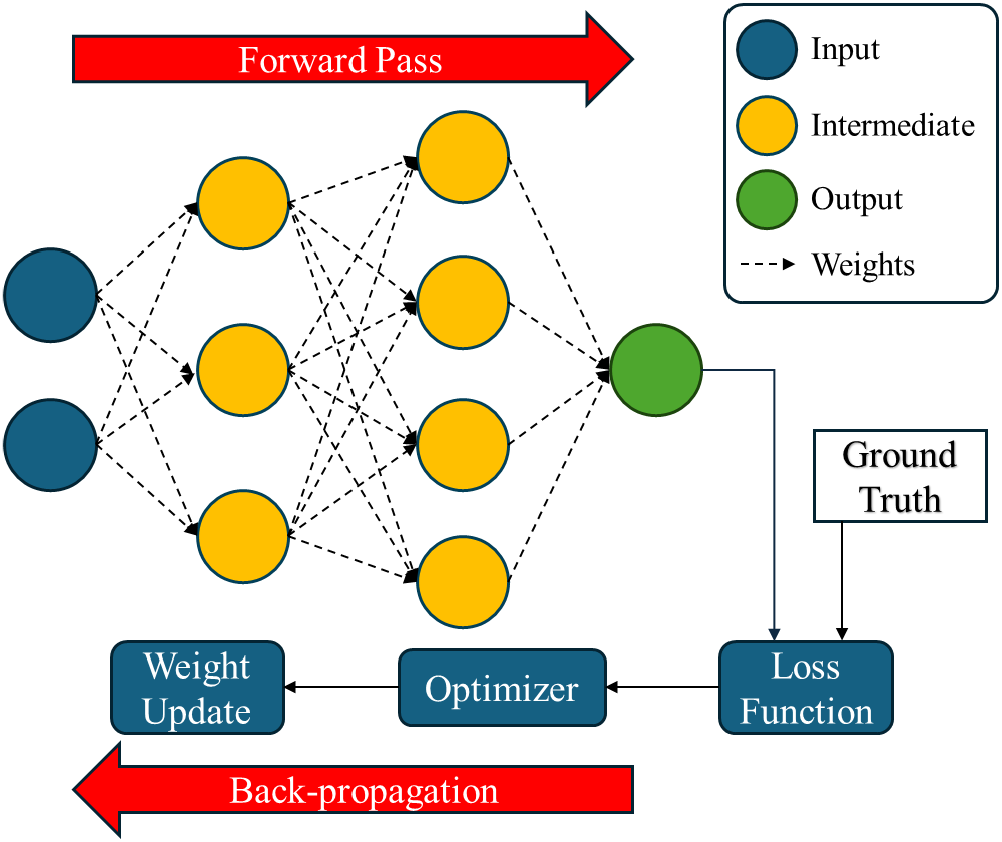
A diagram of an ANN model. The input data is fed into the input layer which is then passed to the intermediate or hidden layers and output at the output layer. The model predictions are then used to compute the loss value to update the weight parameters using an optimizer.

### 1.3 Taxonomy of Deep Learning Tasks

Figure 2 summarizes the different types of deep learning tasks along with a few examples of dental applications. Here we focus on two types of deep learning: supervised and unsupervised learning. We also outline the evaluation metrics used to measure the performance of the deep learning models for each task in the following subsections.

**Figure 2:**
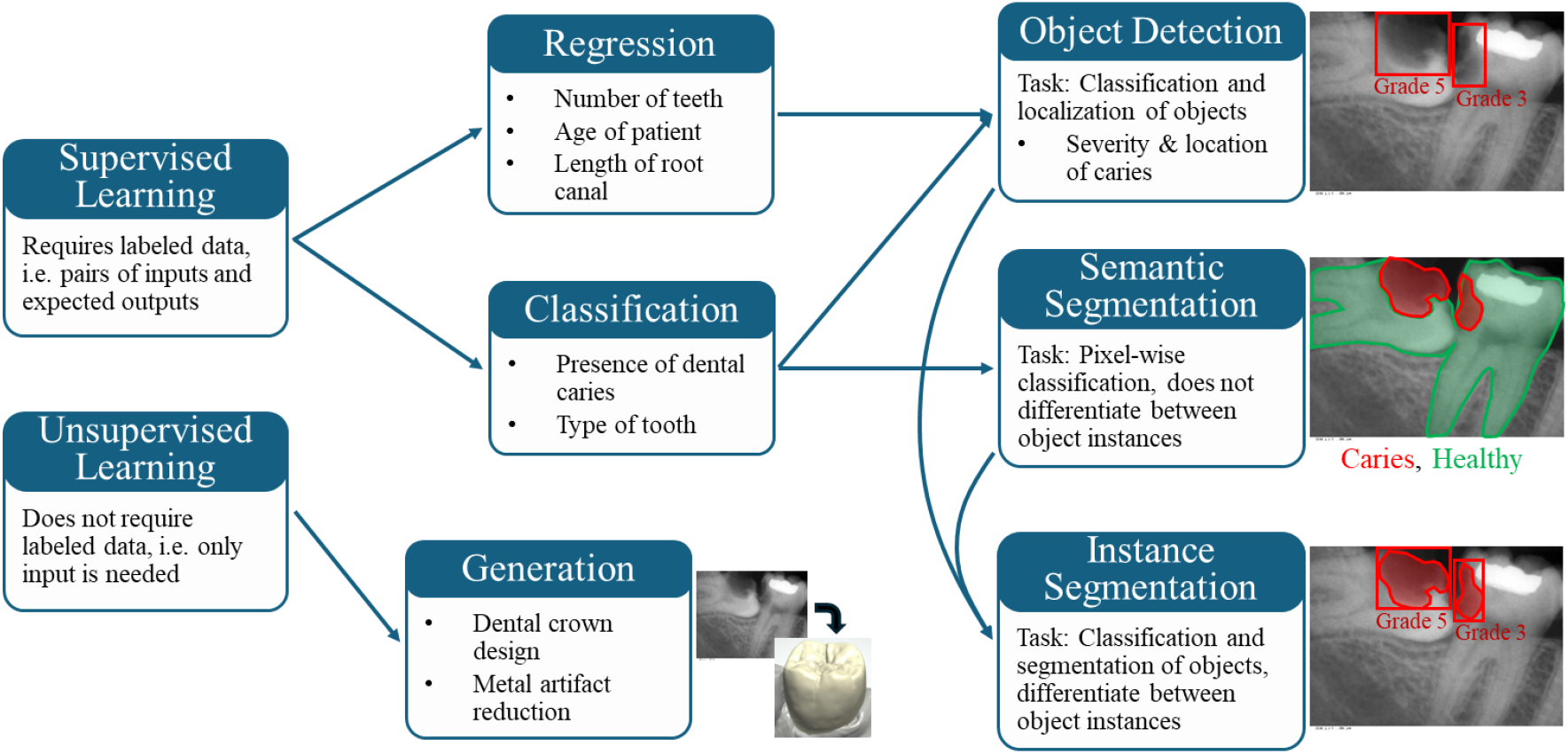
Taxonomy of deep learning tasks along with examples of dentistry tasks. Note that the object detection and instance segmentation tasks expect separate predictions for each object instance while semantic segmentation expects only one prediction mask per class label. The radiograph image and the example outputs are meant as a demonstration only and do not represent the ground truth.

#### 1.3.1 Classification

Classification is a supervised learning task that maps input into a class label. In a c-class classification problem, an ANN with c nodes in the output layer and a SoftMax function predict class probabilities. In the classification task, cross-entropy loss is used to learn the model’s weight. Model performance is evaluated using true positives (TP), false positives (FP), false negatives (FN), and true negatives (TN), typically shown in a confusion matrix. In addition, we can also quantitatively compute several metrics based on the TP, FP, FN, and TN, such as accuracy (ACC), sensitivity (SEN), specificity (SPE), positive predictive value (PPV), negative predictive value (NPV), false positive rate (FPR), and F1 score (Figure 3).

**Figure 3:**
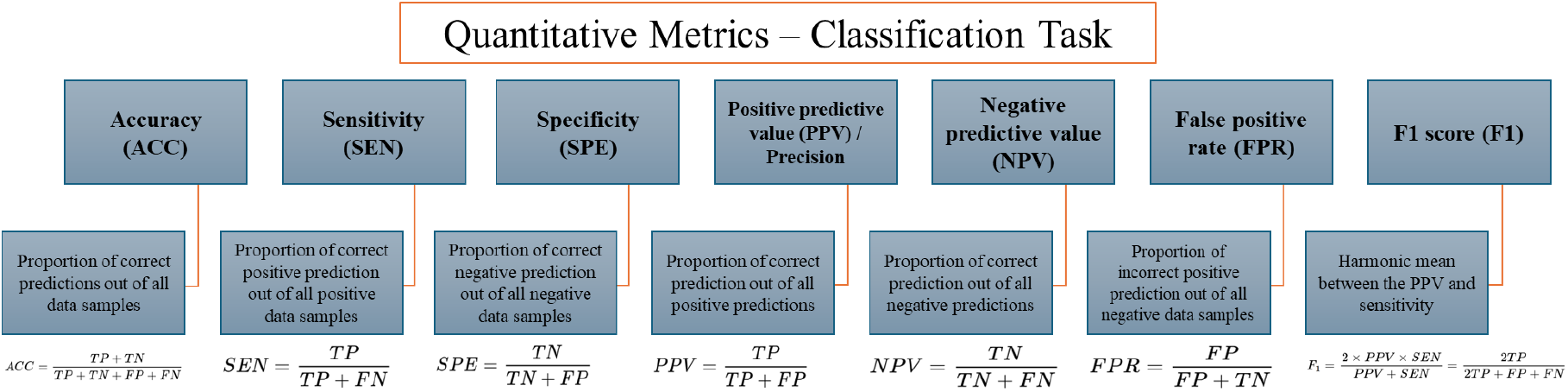
Definition of quantitative metrics for the classification task.

The receiver operating characteristic (ROC) curve plots sensitivity against FPR at various confidence thresholds, with the area under the curve (AUC) measuring overall model performance. It is important to calibrate the model’s sensitivity and specificity by adjusting the confidence threshold. For instance, in a classification model for dental caries, setting a high confidence threshold increases specificity, reducing the likelihood of disease-free patients undergoing unnecessary procedures. Additionally, in cases of class imbalance, using multiple evaluation metrics is vital to prevent misleading performance assessments that disproportionately reflect performance in the majority class.

#### 1.3.2 Regression

Regression aims to learn the relationship between the input features and a continuous target variable. Unlike classification, which produces discrete outputs, regression predicts values in a continuous space (values that can take on any value within a range). Regression can be modeled as an ANN with one or more output nodes, incorporating an activation function to constrain the output range. Common loss functions used for training include mean absolute error loss and mean squared error loss. An example in dentistry is predicting patients’ age using panoramic radiographs. To evaluate a regression model qualitatively, a residual plot can be generated which shows model predictions against the residuals (differences between predictions and actual values). Moreover, several quantitative metrics are used to assess regression performance, including mean error (ME), mean absolute error (MAE), mean squared error (MSE), root mean squared error (RMSE), and R-squared (R^2^) (Figure 4).

**Figure 4:**
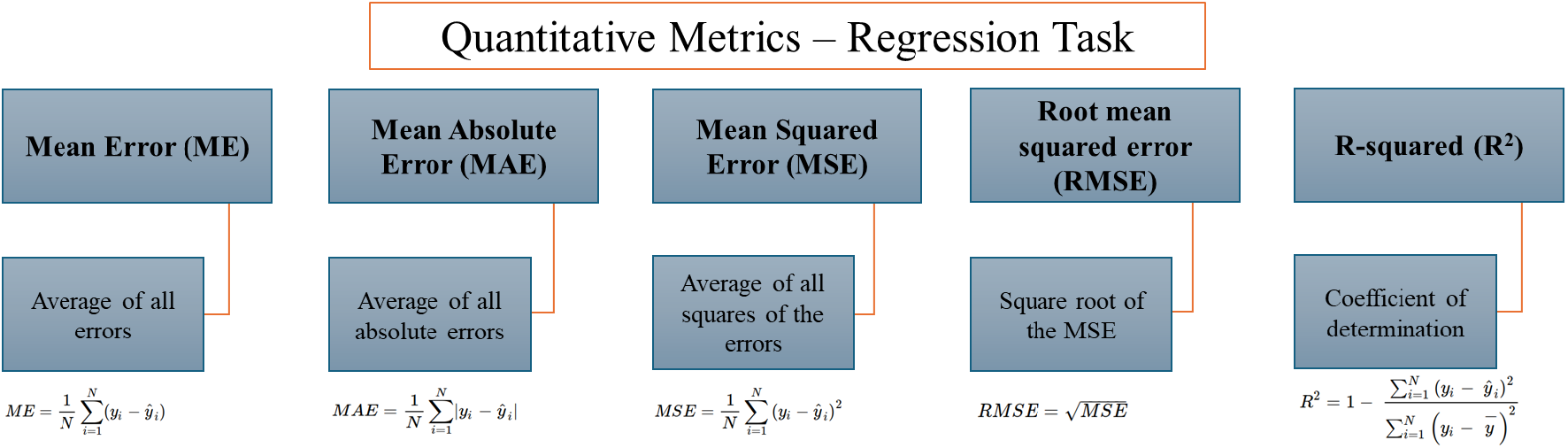
Definition of quantitative metrics for the regression task. Here, N is the number of data samples,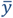 denotes the mean of all ground truth data samples, while y_i_ and ŷ_i_ are the ground truth and predicted value of the i-th data sample, respectively.

#### 1.3.3 Object Detection

Object detection combines both classification and regression tasks to detect instances of visual objects in an image, including their location and class label. Formally, this involves an ANN outputting *M* object predictions, each represented by class probabilities (classification) and object locations (regression). The location of an object is typically defined as the smallest rectangular bounding box enclosing it. Object detection algorithms can be broadly categorized into two types: (a) One-stage detectors, such as YOLO, which prioritize speed; (b) Two-stage detectors, such as Faster R-CNN, which focus on achieving higher accuracy. Before discussing evaluation metrics, it is important to understand the concept of intersection over union (IoU). IoU, a.k.a. Jaccard’s Index, measures the overlap between two bounding boxes by dividing their intersection by their union. It is expressed as follows:

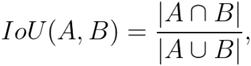

where *A* and *B* represent the set of all pixels belonging to bounding boxes A and B, respectively. IoU measures how well the model localizes objects in an image, with higher scores indicating better accuracy. A common approach in dental research is to set an IoU threshold; predictions above this threshold are considered true positives. Using this threshold, classification metrics can be calculated (Section 1.3.1), though specificity and negative predictive value cannot be determined unless negative labels are present, *e*.*g*. bounding boxes of non-carious tooth in a carious tooth object detection task. Another key metric for evaluating object detectors is mean average precision (AP), *i*.*e*. the area under the precision-recall (PR) curve, showing precision at different recall values across various confidence thresholds. The area is computed by segmenting the recall values of the PR curve into 11 intervals, *i*.*e*.{0,0.1,0.2, …,1},

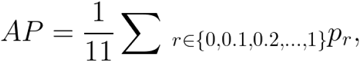

where *p*_*r*_ is the maximum precision obtained at the recall value *r*. In practice, deep learning researchers usually follow the COCO evaluation method and report the mean AP^.5^, AP^.75^ and AP^.5:.95^ computed as the mean AP at IoU thresholds of 0.5, 0.75 and the average of mean AP at 10 IoU thresholds, {0.5, 0.55, …, 0.95}, respectively.

#### 1.3.4 Semantic Segmentation

Semantic segmentation involves assigning a class label to every pixel in an input image, *i*.*e*. pixel-wise c-class classification task. In semantic segmentation, an ANN model outputs a *H* × *W* × *c*matrix representing the class probabilities of each pixel in an input image with a width of *W* and a height of *H*, thus producing c segmentation masks. An example of a deep learning model designed for semantic segmentation tasks is U-Net which is an encoder-decoder-based model widely used in biomedical image segmentation.

Since semantic segmentation is essentially a pixel-level classification task, evaluation metrics defined in Figure 3 can be applied. However, as the number of background (negative class) pixels is typically far greater than the number of pixels for classes of interest (positive class), metrics that use TN can be misleading. Therefore, metrics like the F1 score (or Dice coefficient) are preferred among researchers. Additionally, boundary-based metrics, including the Hausdorff distance (the maximum distance between boundaries) and the average symmetric surface distance (ASSD), can also be used to assess model accuracy.

#### 1.3.5 Instance Segmentation

Instance segmentation aims to predict the segmentation mask for each object in an image, combining object detection and semantic segmentation. For example, we can train a Mask R-CNN model to detect the location and segmentation mask of each tooth in panoramic radiographs. The model is evaluated similarly to an object detection model by calculating the IoU, using the predicted and ground truth masks instead of bounding boxes.

#### 1.3.6 Image Generation

Image generation is an unsupervised task of generating an image given input data. It can be within the same modality (*e*.*g*.image denoising or image super-resolution) or cross-modality (*e*.*g*. generating images from texts). Typically, a generative adversarial network (GAN) is adopted in the image generation task, which consists of two components: a generator neural network and a discriminator neural network. In dentistry, GANs are used to generate dental crown designs from tooth radiographs (8). To evaluate output quality, image generation can be treated as a pixel-wise regression task and reuse the evaluation metrics (Figure 4). Further, the quality of generated images can be assessed with metrics such as peak signal-to-noise ratio (PSNR) or structure similarity index measure (SSIM).

### 1.4 Evaluation Strategy

After training deep learning models on the collected training dataset, the next step is evaluating their performance on the dental task. A common approach is the train/validation/test split: the training set for model learning, the validation set for hyperparameter tuning, and the test set for final performance reporting. However, this method can introduce split bias, especially with small datasets. Alternatively, K-Fold cross-validation divides the dataset into equal parts, training models on all but one part and reporting their average performance. This approach provides a more comprehensive evaluation by aggregating results from distinct models evaluated on the entire dataset.

### 1.5 Modality in Dentistry

Common input modalities in dental tasks include radiographs (bitewing, periapical, occlusal, panoramic, teleradiograph), cone beam computed tomography (CBCT), near-infrared images, and intraoral images. Other modalities include 3D dental models, patient records, and inspection reports *(i*.*e*. text modalities).

## 2 Methods

This survey focuses exclusively on deep learning applications in dentistry, referring readers to earlier reviews for studies using classical machine learning methods (9–11). Following a comprehensive searching and screening process, studies were grouped into two categories: (a) those reusing or slightly modifying existing deep learning models, and (b) those proposing novel deep learning methods. Due to the extensive number of studies, this review focuses on and includes studies in the second group.

### 2.1 Search strategy

We conducted an electronic database search for relevant studies through PubMed, IEEE Xplore, arXiv, and Google Scholar. Furthermore, we recursively searched the reference list of the pre-selected studies that met the inclusion criteria. The last search was performed on the 27^th^ of November 2024, and due to the large number of studies, we only included studies published since 2015. We used a keyword search of “artificial intelligence”, “machine learning”, “deep learning” AND “dentistry”, “dental” with the PubMed query being ‘((“artificial intelligence”[Title] OR “machine learning”[Title] OR “deep learning”[Title]) AND (“dentistry”[Title/Abstract] OR “dental”[Title/Abstract])) AND (2015:2024[pdat])’.

### 2.2 Eligibility criteria

This review exclusively included studies that proposed novel methods utilizing deep learning techniques within the field of dentistry. The screening protocol followed a two-step process: all studies were initially screened by the title and abstract, followed by a full-text review. The exclusion criteria were as follows:

- Studies that are not related to deep learning and dentistry;

- Studies that used existing off-the-shelf deep learning models,

- Studies published in languages other than English;

- Conference abstracts, reviews, expert opinions, editorials, letters, and study protocols.

## 3 Results

Our search strategy yielded 3,030 publications. Additionally, 140 articles were identified manually through a recursive search on the references of screened records. After removing duplicates, 2,473 articles remained. In the first screening, 2,086 articles were excluded based on the titles and abstracts. The remaining 527 full-text articles were assessed for eligibility and 91 articles met the inclusion criteria. Figure 5 outlines the selection process.

**Figure 5:**
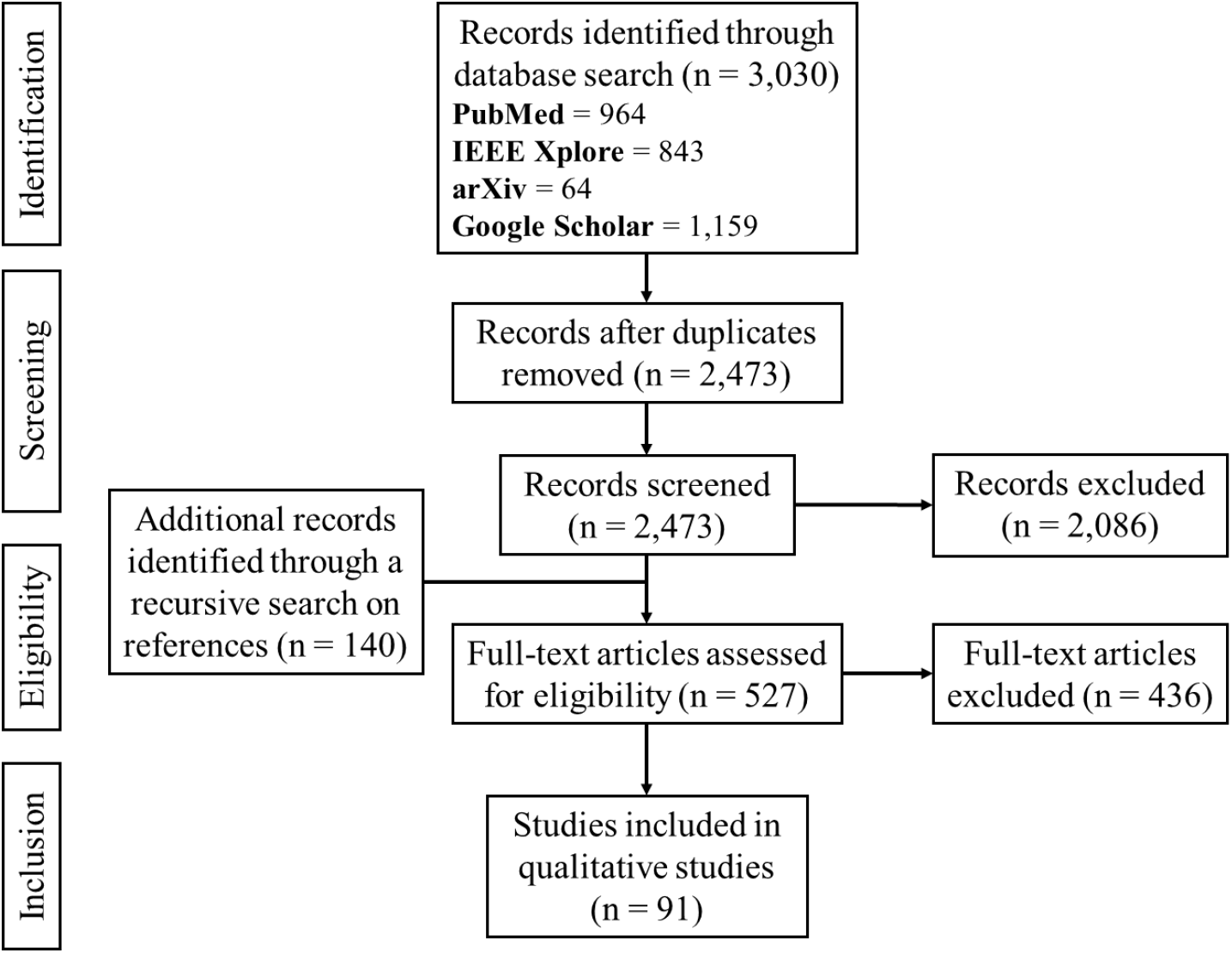
PRISMA flow diagram for the systematic review detailing the database searches, the number of abstracts screened and the full texts assessed.

The included studies are grouped into six domains: (a) Tooth segmentation and classification, (b) Tooth numbering, (c) Cephalometric landmarks, (d) Dental diseases diagnosis, (e) Prosthodontics and Implantology, and (f) Other applications (See Appendix Tables **Error! Reference source not found**.-**Error! Reference source not found**. for a comprehensive analysis for each domain). Panoramic radiography (n=24) was the most used modality, followed by CBCT (n=23) and 3D Intraoral (n=14). Most studies investigate the deep learning tasks of semantic segmentation (n=39), instance segmentation (n=17), classification (n=15), object detection (n=9), and generation (n=9). Validation was typically performed with a train-validation-test split (n=74).

### 3.1 Tooth Segmentation and Tooth Classification

Tooth segmentation and classification are essential for generating detailed dental reports. Xu, Liu (12) used a CNN to segment all teeth in a 3D dental model and another CNN for inter-tooth segmentation. Zheng, Chen (13) introduced a two-branch graph neural network to predict tooth class probabilities of 3D models. Traditional segmentation methods such as the level set algorithm (14) have also been integrated into deep learning methods for better segmentation based on classic methods. Studies are emerging on detecting tooth regions in CBCT scans. Cui, Li (15) trained a 3D Mask R-CNN to perform segmentation and detection, while Chung, Lee (16) used a 3D Faster R-CNN for detection and a UNet for segmentation. Several studies (17, 18) first regressed the centroid coordinates of each tooth, then employed PointNet++ and a CNN to predict the tooth segmentation mask and bounding box around the predicted centroid. The aforementioned studies designed multi-stage frameworks which improve the explainability of the deep learning methods by breaking the final objective into intermediary tasks, they are more time-consuming and susceptible to error accumulation. Besides, Tan, Zhu (19) introduced the U-Mamba2-SSL that progressively utilizes unlabeled CBCT scans to for semi-supervised learning of tooth and pulp segmentation through pre-training, consistency regularization training and pseudo labeling.

Some studies proposed modifying existing model architectures to improve model performance (14, 20, 21) by adding attention modules, multi-scale spatial pyramid pooling, squeeze and excitation modules. However, these approaches simply introduce or combine recent deep learning techniques into existing ones and often lack prior knowledge about tooth structure. To address this, Wirtz, Mirashi (22) used a UNet to create segmentation masks by initializing a shape model based on each tooth’s spatial relationships within the oral cavity. Using prior knowledge, such as the relational position of the crown and root of the tooth, Vinayahalingam, Goey (23) designed a rule-based post-processing to refine Mask R-CNN predictions, while Xie, Yang (24) proposed an ellipse detection method and level-set-based curvature model to approximate tooth boundaries from box-level tooth predictions. Tan, Zhu (25) incorporates several dental domain knowledge such as anatomy structural similarity, relative structural differences and left-right anatomy symmetry to significantly enhance the multi-anatomy segmentation performance of U-Mamba2 in CBCT. The incorporation of these dental domain knowledge makes it easier to explain the underlying mechanism of the proposed deep learning models, improving its explainability and trustworthiness when compared to other approaches.

### 3.2 Tooth Numbering

A natural extension to the task of tooth segmentation and classification is the tooth numbering, which usually follows the FDI notation. Firstly, Zanjani, Anssari Moin (26) used PointCNN to perform 3D segmentation of intraoral scans, then utilized a discriminator network to penalize unrealistic tooth numbering arrangement predictions by the PointCNN model in an adversarial training setup. Next, Jang, Kim (27) proposed to first project CBCT to a panoramic image for bounding box prediction of tooth numbers, and then segment tooth within the predicted box using a UNet. On the other hand, Hao, Wong (28) proposed to utilize unlabeled panoramic radiographs for semi-supervised learning via a teacher-student knowledge distillation strategy. Although these methods achieved respectable performance, they treat the tooth numbering task as separate detection or segmentation tasks, ignoring their inherent ordinal relationship. Since tooth numbers have a concrete definition, dental notation can be easily determined by its position or the neighboring tooth numbers. Several studies incorporated such prior knowledge to obtain a set of more plausible tooth number predictions. For example, Chen, Zhang (29) introduced a post-processing technique that takes advantage of domain knowledge related to tooth positions and similarity weight between tooth categories to arrange the tooth number predictions. Besides, others applied a calibration and optimization procedure to select the best combination of predicted tooth numbers based on the spatial relationship between the predicted bounding boxes (30–32). Although the incorporation of tooth numbers’ spatial relationships generally improves model performance and explainability, it requires meticulously translating the domain knowledge into mathematical constraints or functions. Hence, there exist great research opportunities for automating the process of rendering dentistry domain knowledge into deep learning models.

### 3.3 Cephalometric Landmarks

Several researchers have developed multi-stage detection frameworks for the prediction of cephalometric landmarks of the facial skeleton and skull base. Torosdagli, Liberton (33) first segmented the mandibular area before predicting the mandibular landmarks, while Zeng, Yan (34) proposed a three-stage cascaded CNN to segment the lateral face in cephalometric radiographs, generate landmarks proposals and refine them using 19 CNNs. Furthermore, Chen, Ma (35) developed a framework that detects landmarks from down-sampled CBCT volumes and uses ConvLSTM to refine predictions iteratively. On the other hand, Hwang, Kang (36) introduced SinusC-Net to predict landmark locations from CBCT and used the distances between predicted landmarks for ABC sinus augmentation classification. Lastly, Zhang, Zhao (37) adopted Faster R-CNN model to first detect the location of facial structures (*i*.*e*. ear, eye, jaw) before using ViTPose model to predict the cephalometric landmarks on the cropped images. Many of the studies discussed utilizing a multi-stage framework where different deep learning models each specializing in specific tasks. However, model errors often accumulate and increase across the various stages as the results of the preceding stage are fed into the subsequent deep learning model.

### 3.4 Dental Diseases Diagnosis

Dental caries and periodontal diseases are two of the most common oral diseases. Early detection of caries in imagery is often inconsistent due to high inter-observer variability and low sensitivity (5). To address these challenges, several deep learning methods have been explored. Some models incorporate mathematical constraints, such as ensuring caries regions overlap with tooth and extracting crown regions. Choi, Eun (38) proposed adjusting predicted caries probabilities based on its distance from the crown region, The proposed method, however, requires an extra AI model to perform segmentation of the tooth crown which in turn necessitates the annotation of crown region segmentation mask for model training. Besides that, research also focuses on optimizing deep learning architectures for better caries detection. Haghanifar, Majdabadi (39) suggested using a two-layer capsule network to enhance classification performance. Jiang, Zhang (40) integrated a multi-head attention mechanism into YOLOv5 and ResNet18, while Zhu, Cao (41) used a Res2Net backbone with a full-scale axial attention module on a UNet to propagate information across the image. (42) incorporated Mamba, a selective state space model into UNet to enhance calculus and gingivitis segmentation. Like Section 3.1, these studies only integrate recent deep learning techniques into older ones and do not consider domain knowledge related to dental caries in their modeling approaches.

For periodontal tissues, several deep learning methods have been proposed for detecting alveolar bone loss (ABL).Kim, Lee (43) created a four-stage CNN framework to segment tooth regions and ABL lesions, followed by tooth-level ABL classification for premolars and molars. Other approaches include heuristic methods that utilized bone levels, the cementoenamel junction (CEJ), and tooth segmentation masks (44); and key points of teeth in a periapical radiograph (45). These appraoches utilized domain knowledge where the severity of ABL is determined based on the predicted landmarks such as the CEJ line and the bone-to-tooth intersection line. Lastly, Park, Erkinov (46) proposed using two parallel 1D convolutional layers to enhance periodontal disease classification in tooth radiographs but the proposed method did not bring much innovation other than the use of parallel convolution layers in CNN.

### 3.5 Prosthodontics and Implantology

AI algorithms have revolutionized prosthodontics and implantology fields by enhancing the performance of digital planning pathways. Tian, Wang (8) introduced a two-stage GAN model to reconstruct dental crowns, focusing on recovering the occlusion spatial relationship and generating fine-grained occlusal surface features closer to the true target. UNet models have been explored for error compensation in 3D printing to enhance accuracy (47), taking dental model as input and outputs the scanned 3D-printed dental crown model. Lastly, Yang, Xie (48) proposed TCSIoT, a multimodal model guided by text descriptions of tooth region and CT slices to predict dental implant locations. Considering the broad potential applications of AI in prosthodontic and implantology, the studies discussed in this section each focus on conventional deep learning techniques for one specific task. There exists a research gap in utilizing recent large models in clinical tasks such as dental implant suggestions from intraoral scans, treatment and surgery planning, and much more.

### 3.6 Other Applications

Several studies focus on the segmentation and regression of the mandible and its landmarks. Lian, Wang (49) designed DTNet with local and global branches to segment the lower jaw and localize mandibular landmarks. Some works modified existing deep learning models to segment the mandible in CBCT (50, 51). Cipriano, Allegretti (52) proposed processing cropped CBCT subvolumes, incorporating their global position to translate 2D labels into 3D inferior alveolar nerve annotations. Torosdagli, Anwar (53) proposed a relational reasoning network that effectively learns the relationships among mandibular landmarks.

Other than that, Li, Pang (54) adopted DeepLabV3 trained with a small sample of calculus images for semantic segmentation of dental calculus and tooth. Additionally, Li, Zeng (55) used HRNet and applied polynomial curve fitting to obtain tooth apical boundary for automatic evaluation of root canal filling (*i*.*e*. over-filling, correct-filling, under-filling). (56) proposed FPA-CNN which combines feature pyramid modules and a saliency-driven soft attention module to detect vertical root fractures. Moreover, Lai, Fan (57) proposed LCANet with a channel attention module to enhance human identification accuracy in panoramic radiographs. Finally, Du, Liang (58) proposed a UNet model for metal artifact reduction in CT slices, trained with simulated artifacts by aligning features between real and simulated images.

## 4 Discussion

### 4.1 Incorporation of Dental Domain Knowledge

The intersection of deep learning and dentistry has evolved in several directions due to significant efforts from both research communities. Many studies focus on creating specialized frameworks for multiple dental tasks or innovating deep learning architectures. While these advancements improve performance, they often lack grounding in dental domain knowledge, with few efforts made to integrate such expertise into deep learning designs. Notable exceptions include the incorporation of the structure and shape of tooth into the tooth segmentation task (22, 24, 25) and the use of the relational position and tooth number of neighboring teeth for tooth numbering tasks (30–32). These approaches have resulted in more reasonable diagnosis, better alignment with dental knowledge, and improved performance, making AI diagnosis more trustworthy and explainable for dental practitioners. Thus, collaboration between deep learning and dental researchers to integrate dental domain knowledge into models is crucial and holds significant potential for future research.

### 4.2 Data and Model Uncertainty

Most existing studies reviewed assume no data uncertainty, using available labels directly for training. However, this assumption overlooks the complexity and uncertainty present in many dental tasks (59). As high interobserver variability is observed even among experienced dentists when diagnosing dental diseases, the ‘ground truth’ (typically annotated by one dentist or a consensus of multiple dentists) in dental tasks is not absolute and should be aggregated with care (60) or treated as uncertain data. Furthermore, recent studies have shown that deep learning models struggle to generalize under domain shifts, assigning high confidence to incorrect predictions when presented with out-of-distribution samples (61). In dentistry, domain shifts frequently arise due to variations in patient demographics, imaging protocols, and equipment, which can lead to misdiagnoses and unnecessary procedures with associated costs and risks. Therefore, understanding and capturing model uncertainty is essential, enabling users to evaluate whether to trust the AI model or defer to a human expert.

### 4.3 Large Public Datasets and Large Models

Another significant limitation is the scarcity of publicly available dental datasets. Most articles included in this review do not share their datasets due to factors including consent, rights to share, and privacy protection. Publicly available, properly anonymized datasets are essential for fair evaluation and reproducibility. We have identified 44 dental datasets, with most targeting the semantic segmentation of caries or tooth (Appendix Table 7). These datasets are generally small, with the largest containing just 4,288 labeled bitewing images, a stark contrast to large-scale datasets commonly used in deep learning, such as ImageNet (over 14 million images) and MS COCO (328 thousand images). In the era of large models, these limitations can hinder the development of large, powerful models in dentistry, and the utility of self-supervised or semi-supervised approaches (19, 28, 62). Consequently, foundation models for various applications in dentistry remain unexplored.

### 4.4 Efficient Deep Learning Models

The efficiency of training and inference pipelines is a critical consideration when designing deep learning frameworks. However, many studies prioritize performance over efficiency. For instance, maintaining multiple deep learning models often increases computational costs and reduces pipeline efficiency (16–18). Additionally, when models are applied sequentially, errors can accumulate, degrading overall performance. In contrast, efficient algorithms with negligible drop in model performance can improve scalability with data and are beneficial in environments with limited computational budgets and resources such as dental practices.

### 4.5 Evaluation in Simulated and Real-world Settings

Recent studies have begun to investigate the diagnostic performance and economic benefits of incorporating AI in both simulated and real-world settings. Although randomized controlled trials (RCTs) are considered the gold standard for evaluating AI interventions (63), there is a limited number of peer-reviewed RCT in this field. In an RCT by Cantu, Gehrung (64), the AI-assisted system demonstrated a higher mean sensitivity for caries segmentation compared to dentists (0.75 vs 0.36), but slightly lower specificity (0.83 vs 0.91). However, the same group of four dentists annotated both the training and testing data, which resulted in potential correlations between annotations sets, despite two additional dentists classifying the test annotations. A follow-up RCT, (65) involving a reference test established similarly with an additional independent expert, found that AI improved sensitivity (0.81 vs 0.72) while maintaining similar specificity (0.97). A cost-effectiveness analysis of AI (66) revealed that despite AI’s higher accuracy it did not offer t greater cost-effectiveness, with costs estimated using the German public and private fee catalogues. In another study (67), a CNN-based system for periodontal detection on panoramic radiographs was found to perform similarly to experts and outperformed general dentists. These studies provide some of the most comprehensive evaluations of dental AI-assisted systems with robust statistical results. Nonetheless, further independent assessments and comparisons of available tools using objective datasets with ground truth reference annotations would be highly valuable for advancing the field.

## 5 Conclusion

Our study first clarified key terminologies and concepts in AI, then presented a comprehensive analysis of the 91 articles included. The findings highlighted the transformative potential of AI in revolutionizing dentistry, with recent novel advancements in deep learning demonstrating great potential in various domains such as diagnosis, prosthodontics and implantology. However, the review also identified several challenges to fully realize the potential benefits, which necessitate further research in multiple aspects including integrating dental knowledge, quantifying uncertainty, utilizing large datasets, developing efficient deep learning pipelines, and comprehensive evaluation in simulated and real-world environments. Addressing these challenges in future research are essential for empowering dental professionals to leverage deep learning and AI effectively to transform dentistry into a more precise, efficient, and patient-centered field, ultimately leading to enhanced clinical outcome and patient care.

## Declarations

### Clinical trial number

Clinical trial number: not applicable.

### Ethics approval and consent to participate

Not applicable.

### Consent for publication

Not applicable.

### Data availability

Data sharing is not applicable to this article as no datasets were generated or analyzed during the current study.

## Acknowledgements

This work was supported by the authors’ university department, which provided the authors with the necessary environment and resources for this work.

## Authors’ contributions

Z.Q.T. contributed to the design, data acquisition, analysis and interpretation, drafted and critically revised the manuscript. M.G.R. contributed to the data analysis and interpretation, drafted and critically revised the manuscript. O.A. contributed to the conception, data interpretation, and critically revised the manuscript. Y.L. contributed to the conception and design, data interpretation, and critically revised the manuscript. All authors read and approved the final manuscript.

