## Supplementary Material for "Deep Learning in Dentistry: A Systematic Review from an AI Researcher Viewpoint"

Zhi Qin Tan, Marina Guimaraes Roscoe, Owen Addison, Yunpeng Li

Centre for Oral, Clinical & Translational Sciences

Faculty of Dentistry, Oral & Craniofacial Sciences

King’s College London, London, UK

Appendix Table 1: Studies that utilize deep learning (DL) in tasks related to tooth segmentation and classification (excluding tasks of tooth numbering).

| **Study** | **Method** | **Open Source** | **DL Task** | **Target Problem** | **Input Modality** | **Dataset Size** | **Dentistry Domain Knowledge** | **Multi-Stage DL** | **Validation Method** | **Metrics** |
| --- | --- | --- | --- | --- | --- | --- | --- | --- | --- | --- |
| Wirtz et al. (2018) | UNet+Coupled Shape Model | N | Instance Segmentation | Tooth segmentation | Panoramic | 10/14 | Y | Y | Train/Val/Test split | Accuracy, Specificity, Sensitivity, PPV, DICE |
| Xu et al. (2019) | CNN | Y | Instance Segmentation | Tooth segmentation | 3D dental model | 1,000/50/150 | N | Y | Train/Val/Test split | Accuracy, ME |
| Tian et al. (2019) | Multi-branch  CNN | N | Instance Segmentation | Tooth segmentation | 3D dental model | 500/50/50 | N | Y | Train/Val/Test split | Accuracy |
| Gou et al. (2019) | UNet | N | Semantic Segmentation | Tooth segmentation | CT slices | 300/100 | N | N | Train/Val/Test split | Accuracy |
| Cui et al. (2019) | ToothNet | N | Semantic Segmentation | Tooth segmentation | CBCT | 12/8 | Y | Y | Train/Val/Test split | Accuracy, DICE |
| Chen et al. (2020) | UNet+MWT | N | Instance Segmentation | Tooth segmentation | CBCT | 20/5 | N | Y | Train/Val/Test split | DICE, IoU, RVD, ASSD |
| Lian et al. (2020b) | MeshSegNet | N | Instance Segmentation | Tooth segmentation | 3D Intraoral | 30 | N | N | 5-Fold | Sensitivity, PPV, DICE |
| Chung et al. (2020) | Faster R-CNN + UNet | N | Instance Segmentation | Tooth segmentation | CBCT | 50/25 | N | Y | Train/Val/Test split | Sensitivity, PPV,  DICE, IoU, AP*^.^*^5^,  Hausdorff distance, ASSD |
| Zhao et al. (2020) | TSASNet | N | Semantic Segmentation | Tooth segmentation | Panoramic | 1,200/150/150 | N | Y | Train/Val/Test split | Accuracy, Specificity, Sensitivity, PPV, DICE |
| Vinayahalingam et al. (2021) | Mask R-CNN | N | Instance Segmentation | Tooth segmentation | Panoramic | 1,600/200/200 | Y | N | Train/Val/Test split | Sensitivity, PPV, DICE |
| Chung et al. (2021) | CNN | N | Object Detection | Teeth detection | Panoramic | 574/162/82 | N | Y | Train/Val/Test split | Sensitivity, PPV,  ROC, IoU, AP*^.^*^5^, AP*^.^*^75^,  AP*.*5:*.*95 |
| Zanjani et al. (2021) | Mask-MCNet | N | Instance Segmentation | Tooth segmentation | 3D Intraoral | 168 | N | N | 5-Fold | IoU, AP*.*25, AR*.*25 |
| Chen et al. (2021) | MSLPNet | N | Semantic Segmentation | Tooth segmentation | Panoramic | 1,200/150/150 | N | N | Train/Val/Test split | Accuracy, Specificity, Sensitivity, PPV, DICE |
| Nishitani et al. (2021) | UNet | N | Semantic Segmentation | Tooth segmentation | Panoramic | 82/20/60 | Y | N | Train/Val/Test split | DICE, IoU |
| Cui et al. (2021) | TSegNet | N | Instance Segmentation | Tooth segmentation | 3D Intraoral | 1,500/100/400 | N | Y | Train/Val/Test split | DICE |
| Gao et al. (2022) | Grouped  Bottleneck  Transformer | Y | Classification | Teeth classification | CBCT | 450/104 cropped tooth images | N | N | Train/Val/Test split | Accuracy, ROC |
| Wu et al. (2022) | MeshSegNet | N | Semantic Segmentation, Regression | Tooth segmentation and landmarks | 3D Intraoral | 65/15/20 | N | Y | Train/Val/Test split | Sensitivity, PPV, DICE, Hausdorff distance, MAE |
| Qiu et al. (2022) | DArch | N | Instance Segmentation | Tooth segmentation | 3D dental model | 3973/800 | Y | Y | Train/Val/Test split | Accuracy, Sensitivity,  DICE, IoU |
| Zhao et al. (2022) | TSGCN | N | Semantic Segmentation | Tooth segmentation | 3D Intraoral | 64/16 | N | N | Train/Val/Test split | Accuracy, IoU |
| Liu et al. (2023b) | STSNet | N | Semantic Segmentation | Tooth segmentation | 3D Intraoral | 12,000 unlabeled +600/200/200 labeled | N | N | Train/Val/Test split | Accuracy, DICE, IoU |
| Hou et al. (2023) | Teeth UNet | N | Semantic Segmentation | Tooth segmentation | Panoramic | 1,200/150/150 | N | N | Train/Val/Test split | Accuracy, Sensitivity,  PPV, DICE, IoU |
| Zheng et al. (2023) | TeethGNN | N | Semantic Segmentation | Tooth segmentation | 3D dental model | 3,456/372 | N | N | Train/Val/Test split | Accuracy, IoU |
| Xie et al. (2023) | WITS | Y | Instance Segmentation | Tooth segmentation | CBCT | 7/3 | Y | N | Train/Val/Test split | PPV, DICE, IoU |
| Lin et al. (2024) | DBGANet | Y | Semantic Segmentation | Tooth segmentation | 3D dental models | 40 | N | N | Train/Val/Test split | Sensitivity, Precision, DICE |
| Li et al. (2024b) | HiCA | Y | Semantic Segmentation | Tooth segmentation | 3D Intraoral | 76 | N | N | 5-Fold | Accuracy, DICE |
| Li et al. (2024a) | UNet w/ multi-scale attention | N | Semantic Segmentation | Tooth segmentation | CBCT | 2260/588 | Y | Y | Train/Val/Test split | Sensitivity, Precision, DICE, IoU, ASSD |
| Zhou et al. (2024) | WTNet | N | Semantic Segmentation | Wisdom tooth segmentation | CBCT | 133 | N | Y | 5-Fold | Accuracy, IoU, Hausdorff distance, ASSD |
| Chen et al. (2024a) | nnUNet | N | Semantic Segmentation | Tooth segmentation | CBCT, 3D Intraoral | 1417 CBCT, 1801 3D Intraoral | N | Y | Train/Val/Test split | Sensitivty, DICE, ASSD |
| Almalki and Latecki (2024) | DentalMAE | N | Semantic Segmentation | Tooth segmentation | 3D Intraoral | 1200/600 | N | N | Train/Val/Test split | Accuracy, sensitivity, Precision, DICE |
| Li et al. (2024c) | THISNet | N | Instance Segmentation | Tooth segmentation | 3D Intraoral | 1200/600 | N | N | Train/Val/Test split | Accuracy, IoU |
| Ahn and Cho (2024) | WCTN | N | Instance Segmentation | Tooth segmentation | 3D Intraoral | 1587 | N | N | 5-Fold | Accuracy, IoU |
| Ding et al. (2024) | TP-Net | N | Regression | Tooth pose | 3D Intraoral | 288864/69617 | N | N | Train/Val/Test split | MAE |
| Ong et al. (2024) | YOLOv5, UNet, EfficientNet | N | Object Detection, Semantic Segmentation, Classification | Tooth segmentation and classification | Panoramic | 5133 | N | Y | Train/Val/Test split | Sensitivity, Precision, F1, AP |
| Tan et al. (2025a) | U-Mamba2 | Y | Semantic Segmentation | Tooth segmentation | CBCT | 532 | Y | N | Train/Val/Test split | DICE, Hausdorff distance |
| Tan et al. (2025b) | U-Mamba2-SSL | Y | Semantic Segmentation | Tooth segmentation | CBCT | 30 labeled + 300 unlabeled | Y | N | Train/Val/Test split | Accuracy, DICE, IoU |

Appendix Table 2: Studies that utilize deep learning in tooth numbering tasks.

| **Study** | **Method** | **Open Source** | **DL Task** | **Target Problem** | **Input Modality** | **Dataset Size** | **Dentistry Domain Knowledge** | **Multi-Stage DL** | **Validation Method** | **Metrics** |
| --- | --- | --- | --- | --- | --- | --- | --- | --- | --- | --- |
| Zhang et al. (2018) | Combination of CNNs | N | Object Detection | Tooth numbering | Periapical | 800/200 | Y | Y | Train/Val/Test split | Sensitivity, PPV, F1 |
| Chen et al. (2019) | Faster R-CNN | N | Object Detection | Tooth numbering | Periapical | 800/200/250 | Y | N | Train/Val/Test split | Sensitivity, PPV, IoU |
| Zanjani et al. (2019) | PointCNN | N | Semantic Segmentation | Tooth numbering | 3D Intraoral | 120 | N | N | 5-Fold | Sensitivity, PPV, IoU |
| Li et al. (2020a) | Mask R-CNN | N | Instance Segmentation, Classification | Tooth numbering | Panoramic | 302 | Y | Y | 3-Fold | Accuracy, F1, DICE, AP*^.^*^5:^*^.^*^95^ |
| Mahdi et al. (2020) | Faster R-CNN | N | Object Detection | Tooth numbering | Panoramic | 900/100 | Y | N | Train/Val/Test split | Sensitivity, PPV, F1,  IoU, AP*^.^*^5^ |
| Hsu and Wang (2021) | UNet +  Mask R-CNN | Y | Instance Segmentation, Semantic Segmentation | Tooth numbering | Panoramic | 1,200/300 | Y | Y | Train/Val/Test split | Sensitivity, PPV, F1,  ROC, AP^0^, AP*^.^*^5^, AP*^.^*^7^ |
| Jang et al. (2022) | Hierarchical multi-step model | N | Instance Segmentation | Tooth numbering | CBCT | 66/31 | N | Y | Train/Val/Test split | Sensitivity, PPV, DICE, Hausdorff distance,  ASSD |
| Zhang et al. (2022b) | Relation-based  CNN | N | Object Detection | Tooth numbering | Periapical | 1,125/125 | Y | N | Train/Val/Test split | Sensitivity, PPV, F1 |
| Zhang et al. (2022a) | BDUNet | N | Semantic Segmentation | Tooth numbering | Panoramic | 524/275/117 | N | Y | Train/Val/Test split | DICE |
| Shi et al. (2024) | YOLOv3, SOS-Net | N | Object Detection, Classification | Tooth numbering and development stages | Panoramic | 471/68/134 | N | Y | Train/Val/Test split | Sensitivity, Precision, F1, ROC, IoU, AP*^.^*^5^, AP*^.^*^75^ |
| Hao et al. (2024) | SemiTNet | Y | Semantic Segmentation | Tooth numbering | Panoramic | 1398 labeled, 14728 unlabeled | N | N | Train/Val/Test split | Sensitivity, Precision, F1, DICE, IoU |

Appendix Table 3: Studies that utilize deep learning in tasks related to the prediction of cephalometric landmarks of the facial skeleton and skull base.

| **Study** | **Method** | **Open Source** | **DL Task** | **Target problem** | **Input Modality** | **Dataset Size** | **Dentistry Domain Knowledge** | **Multi-Stage DL** | **Validation Method** | **Metrics** |
| --- | --- | --- | --- | --- | --- | --- | --- | --- | --- | --- |
| Qian et al. (2019) | CephaNet | N | Keypoint Detection | Cephalometric landmarks | Cephalometric radiograph | 150/250 | N | N | Train/Val/Test split | Accuracy |
| Torosdagli et al. (2019) | Tiramisu | N | Semantic Segmentation | Facial landmarks | CBCT | 50/48 | N | Y | Train/Val/Test split | IoU, DICE,  Hausdorff distance |
| Zeng et al. (2021) | Cascaded CNN | N | Keypoint Detection | Cephalometric landmarks | Cephalometric radiograph | 150/250 | N | Y | Train/Val/Test split | Accuracy,  Mean radial error |
| Chen et al. (2022) | SA-LSTM | Y | Regression | Cephalometric landmarks | CBCT | 89 | N | Y | 3-Fold | Mean radial error |
| Hwang et al. (2023) | SinusC-Net | N | Keypoint Detection, Classification | Facial landmarks,  Sinus augmentation | CBCT | 72/8/53 | N | Y | Train/Val/Test split | Accuracy, Specificity,  Sensitivity, ROC,  Mean radial error |
| Zhang et al. (2024) | FaLdViT | N | Keypoint Detection | Cephalometric landmarks | Cephalometric radiograph | 300/50/50 | Y | Y | Train/Val/Test split | Accuracy,  Mean radial error |

Appendix Table 4: Studies that utilize deep learning in dental disease diagnosis tasks.

| **Study** | **Method** | **Open Source** | **DL Task** | **Target Problem** | **Input Modality** | **Dataset Size** | **Dentistry Domain Knowledge** | **Multi-Stage DL** | **Validation Method** | **Metrics** |
| --- | --- | --- | --- | --- | --- | --- | --- | --- | --- | --- |
| Choi et al. (2018) | FCN | N | Semantic Segmentation | Caries detection | Periapical | 475 | Y | N | 5-Fold | DICE |
| Haghanifar et al. (2020) | PaxNet | N | Classification | Caries detection | Panoramic | 470/42 | N | Y | Train/Val/Test split | Accuracy, Sensitivity,  PPV, F1 |
| Zheng et al. (2021) | Dense U-Net | N | Semantic Segmentation | Caries, bone, tooth | CBCT | 20 | Y | N | 4-Fold | Sensitivity, PPV,  DICE |
| Jiang et al. (2021) | RDFNet | N | Object Detection | Caries detection | Intraoral | 5,987/1,711/856 | N | N | Train/Val/Test split | Sensitivity, PPV,  AP*.*5 |
| Ying et al. (2022) | Transformer | N | Semantic Segmentation | Caries detection | Periapical | 113/40 | N | N | Train/Val/Test split | Specificity, Sensitivity,  PPV, DICE |
| Jiang et al. (2023) | CariesFG | Y | Classification | Caries detection | Intraoral | 4,495/1,124 tooth images | N | Y | Train/Val/Test split | Accuracy, Specificity, Sensitivity, PPV, F1 |
| Zhu et al. (2023) | CariesNet | N | Semantic Segmentation | Caries detection | Panoramic | 900/135/124 | N | N | Train/Val/Test split | Accuracy, Sensitivity,  PPV, DICE |
| Zhou et al. (2023) | T2S Transformer | N | Classification | Caries detection | Panoramic | 4,833/599/596 cropped tooth images | N | N | Train/Val/Test split | Accuracy, Sensitivity,  PPV, F1, ROC |
| Wang et al. (2023b) | MLUA | Y | Semantic Segmentation | Caries detection | Panoramic | 900/100 | N | N | Train/Val/Test split | Sensitivity, PPV,  DICE |
| Liu et al. (2024b) | ResNet50 w/ spatial attention mechanism | N | Classification | Caries detection | Periapical | 9042/1728/1754 | N | N | Train/Val/Test split | Accuracy, Specificity, Sensitivity, Precision, F1 |
| Liu et al. (2024a) | Oral-Mamba | N | Semantic Segmentation | Caries, gingivitis, calculus | Intraoral | 2019/673/673 | N | N | Train/Val/Test split | Accuracy, Sensitivity, Precision, IoU |
| Kim et al. (2019) | DeNTNet | N | Semantic Segmentation, Classification | Alveolar bone loss detection | Panoramic | 11,189/190/800 | Y | Y | Train/Val/Test split | Specificity, Sensitivity,  PPV, F1, ROC, NPV |
| Chang et al. (2020) | Mask R-CNN | N | Semantic Segmentation, Classification | Alveolar bone loss detection | Panoramic | 340 | Y | N | Train/Val/Test split | Accuracy, DICE, IoU |
| Danks et al. (2021) | Hourglass | Y | Keypoint detection | Tooth landmarks  Periodontal bone loss | Periapical | 340 | Y | N | 3-Fold | MSE |
| Cha et al. (2021) | Mask R-CNN | N | Object Detection, Keypoint detection | Implants,  peri-implant bone loss | Periapical | 508/100/100 cropped tooth images | Y | N | Train/Val/Test split | AP*.*5, AP*.*75, AP*.*5:*.*95,  AR*.*5:*.*95 |
| Park et al. (2023) | ResNet152 w/ parallel convolution | N | Classification | Periodontal diseases | Intraoral | 220 tooth images | N | N | 10-Fold | Accuracy, ROC |
| Xue et al. (2024) | YOLOv8, Mask R-CNN, TransUNet | N | Instance Segmentation | Teeth, Tissue type, Periodontal bone loss | Panoramic | 288/32 | Y | Y | Train/Val/Test split | Accuracy, Sensitivity, Precision, F1 |

Appendix Table 5: Studies that utilize deep learning in prosthodontics and implantology tasks.

| **Study** | **Method** | **Open Source** | **DL Task** | **Target Problem** | **Input Modality** | **Dataset Size** | **Dentistry Domain Knowledge** | **Multi-Stage DL** | **Validation Method** | **Metrics** |
| --- | --- | --- | --- | --- | --- | --- | --- | --- | --- | --- |
| Hwang et al. (2018) | Pix2pix | N | Generation | Dental crown design | Intraoral scan | 1,500/1,570/243 | Y | N | Train/Val/ Test split | Sensitivity, PPV,  F1, IoU, RMSE |
| Shen et al. (2019b) | 3D UNet | N | Generation | Error compensation in 3D printer | 3D crown model | 77 | N | N | Train/Val/ Test split | F1 |
| Shen et al. (2019a) | PredNet + CompNet | N | Generation | Error compensation in 3D printer | 3D crown model | 57/14 | N | Y | Train/Val/ Test split | Sensitivity, PPV, F1 |
| Tian et al. (2021) | DAIS | Y | Generation | Dental inlay restoration | 3D Intraoral | 750/80 | N | N | Train/Val/ Test split | PSNR, SSIM, RMSE, FSIM |
| Tian et al. (2022) | DCPR-GAN | Y | Generation | Dental crown surface reconstruction | 3D Intraoral | 700/80 | N | Y | Train/Val/ Test split | PSNR, SSIM, RMSE, FSIM |
| Shen et al. (2023) | TranSDFNet | N | Generation | Removable partial denture clasps design | 3D dental model | 30/9 | N | N | Train/Val/ Test split | F1, IoU, Earth mover distance |
| Yang et al. (2023) | TCSIoT | N | Object Detection | Dental implant | CT slices | 3,045 | Y | Y | 10-Fold | F1, AP*.*75 |
| Zhao et al. (2024) | CCMGAN | N | Generation | Dental crown design | 3D dental model | 100 | N | Y | Train/Val/ Test split | Frechet DGCNN distance, Minimum matching distance, Earth mover distance |
| Chen et al. (2024b) | BESO-Net | N | Generation | Prothesis design | Finite element analysis output | 12/2 | N | N | Train/Val/ Test split | ME |

Appendix Table 6: Studies that utilize deep learning in tasks related to bone structure, mandibular, vertical root fracture, dental plaque, etc.

| **Study** | **Method** | **Open Source** | **DL Task** | **Target Problem** | **Input Modality** | **Dataset Size** | **Dentistry Domain Knowledge** | **Multi-Stage DL** | **Validation Method** | **Metrics** |
| --- | --- | --- | --- | --- | --- | --- | --- | --- | --- | --- |
| Zhang et al. (2017) | Stacked UNet | N | Semantic Segmentation, Regression | Craniomaxillofacial bone and landmarks | CBCT | 107 | N | Y | 5-Fold | Sensitivity, PPV,  DICE |
| Lian et al. (2020a) | DTNet | N | Semantic Segmentation | Mandible | CBCT | 120/20 | N | N | Train/Val/Test split | Sensitivity, PPV,  DICE, RMSE |
| Li et al. (2020b) | DeepLabV3 | N | Semantic Segmentation | Dental plaque | Intraoral | 320/287 tooth images | N | Y | Train/Val/Test split | Accuracy, IoU |
| Kong et al. (2020) | EED-Net | N | Semantic Segmentation | Maxillofacial | Panoramic | 2,082/520 | N | N | Train/Val/Test split | Accuracy, IoU,  Hausdorff distance (pixel) |
| Qiu et al. (2021) | C2FSeg | N | Semantic Segmentation | Mandible | CBCT | 38/1/20 | N | Y | Train/Val/Test split | DICE, ASSD,  Hausdorff distance |
| Lai et al. (2021) | LCANet | Y | Classification | Human identification | Panoramic | 22,172/1,168 | N | N | Train/Val/Test split | Accuracy |
| Xu et al. (2021) | FPA-CNN | N | Classification | Vertical root fracture | CBCT slices | 9,219 | N | N | Train/Val/Test split | PR Curve |
| Li et al. (2022) | AGMB-  Transformer | Y | Classification | Root canal therapy | Periapical | 245 cropped tooth w/ root canal therapy | N | Y | 3-Fold | Accuracy, Specificity, Sensitivity, F1, ROC |
| Jeoun et al. (2022) | Canal-Net | N | Semantic Segmentation | Mandibular canal | CBCT | 60/20/20 | N | N | Train/Val/Test split | Sensitivity, PPV,  DICE, IoU, RVD |
| Cipriano et al. (2022) | Positional  PadUNet | Y | Semantic Segmentation | Alveolar nerve | CBCT | 68/8/15 | N | N | Train/Val/Test split | IOU, DICE |
| Kalla et al. (2023) | UNet | N | Regression | Registration of dental images | Panoramic | 116 | N | N | 5-Fold | MSE |
| Torosdagli et al. (2023) | Relational reasoning network | N | Regression | Mandibular landmarks | CBCT | 250 | N | N | 4-Fold | RMSE |
| Du et al. (2023) | GAN | N | Generation | Metal artefact reduction | CT slices | 3,014/861/430 | N | N | Train/Val/Test split | - |

Appendix Table 7: Publicly available datasets grouped by the type of dental task and the input modality. ∗ indicates that the download link is not working at the time of writing.

| **Dataset** | **Year** | **Input Modality** | **DL Task** | **Dental Task** | **Dataset Size** |
| --- | --- | --- | --- | --- | --- |
| Kunt et al. (2023) | 2023 | Bitewing | Object Detection | Caries | 100 |
| teeth_dataset (Pushkara 2020) | 2020 | Intraoral | Classification | Caries | 60/14 |
| teethdecay (Đinh 2022) | 2022 | Intraoral | Classification | Caries | 1,260/294 |
| Liu et al. (2024a) | 2024 | Intraoral | Semantic Segmentation | Caries, Calculus, Gingivitis | 3365 |
| Zhang et al. (2023) | 2023 | Pediatric Panoramic | Semantic Segmentation | Caries | 193 |
| DC1000 (Wang et al. 2023b) | 2023 | Panoramic | Semantic Segmentation | Caries | 593 detailed,  407 rough annotations |
| DENTEX (Hamamci et al. 2023) | 2023 | Panoramic | Object Detection | Quadrant, Tooth numbering, Caries/Lesion/Impacted tooth | 1,005 labeled,  1,571 unlabeled |
| Dental Radiography (Momeni 2023) | 2023 | Panoramic | Object Detection | Caries, Implants, Fillings, Impacted tooth | 1269 |
| Rad et al. (2016) | 2016 | Periapical | Semantic Segmentation | Caries | 120 |
| Rashid et al. (2022) | 2022 | Periapical | Object Detection | Caries | 936 |
| Rashid et al. (2022) | 2022 | Photographic Frontal Tooth Image | Object Detection | Caries | 90 |
| object_detection_on_cavities (Gopinathan 2022) | 2022 | Intraoral | Object Detection | Cavities | 591/123 |
| Calazans et al. (2022) | 2022 | CBCT slices | Classification | Tooth lesion | 1,000 cropped tooth images |
| calculus (Lab 2023) | 2023 | Photographic Frontal Tooth Image | Classification | Calculus | 229 |
| Do et al. (2024) | 2024 | Panoramic | Object Detection | Periodontal lesion | 3926 |
| Dental-Caries-Segmentation (Fatima 2022) | 2022 | Periapical | Instance Segmentation | Periodontal lesion | 458/42 |
| Thalji et al. (2024) | 2024 | Periapical | Instance Segmentation | Periapical lesion | 929 |
| Barot and Suthar (2020) | 2020 | Intraoral | Classification | Oral Cancer | 131 |
| ISBI2015 -Challenge1^∗^ (Wang et al. 2016) | 2016 | Cephalometric | Keypoint Detection | Cephalometric landmarks | 150/250 |
| Wang et al. (2023a) | 2023 | Cephalometric | Keypoint Detection | Cephalometric landmarks | 700 |
| Huang et al. (2024) | 2023 | CBCT  Panoramic  Periapical | Classification | Dental implant of CBCT | 329 CBCT,  8 panoramic,  188 periapical |
| ToothFairy (Cipriano et al. 2022) | 2022 | CBCT | Semantic Segmentation | Alveolar nerve | 68/8/15 |
| PDDCA (Raudaschl et al. 2017) | 2017 | CT | Semantic Segmentation | Mandible | 25/10/5 |
| Abdi et al. (2015) | 2015 | Panoramic | Semantic Segmentation | Mandible | 116/30/95 |
| CTooth (Cui et al. 2022b) | 2022 | CBCT | Semantic Segmentation | Tooth | 22 |
| CTooth+ (Cui et al. 2022a) | 2023 | CBCT | Semantic Segmentation | Tooth | 22 labeled, 146 unlabeled |
| Silva et al. (2018) | 2018 | Panoramic | Semantic Segmentation | Tooth | 1,500 |
| Panoramic Dental Xray Dataset (Brahmi and Jdey 2024) | 2024 | Panoramic | Instance Segmentation | Tooth | 107 |
| 3DTeethSeg’22 (Ben-Hamadou et al. 2023) | 2023 | 3D Intraoral | Semantic Segmentation | Tooth numbering | 1,200 |
| Azleen (2024) | 2024 | Bitewing | Instance Segmentation | Tooth numbering | 3035/847/406 |
| Johamni et al. (2023) | 2023 | Panoramic | Object Detection | Tooth numbering | 1404 |
| Loop (2023) | 2023 | Panoramic | Instance Segmentation | Tooth numbering | 598 |
| Adnan (2023) | 2023 | Panoramic | Instance Segmentation | Tooth numbering | 250 |
| TISI15k-Dataset (Hao et al. 2024) | 2024 | Panoramic | Instance Segmentation | Tooth numbering | 1398 labeled, 14728 unlabeled |
| Costa et al. (2024) | 2024 | Panoramic | Object Detection | Tooth numbering, Caries, Restorations | 936 |
| Teeth3DS+ (Ben-Hamadou et al. 2022) | 2024 | 3D Intraoral | Instance Segmentation | Tooth numbering, Dental landmarks | 1200/600 |
| ISBI2015 -Challenge2^∗^ (Wang et al. 2016) | 2016 | Bitewing | Semantic Segmentation | Tooth structures | 40/80 |
| Cui et al. (2022c) | 2022 | CBCT | Semantic Segmentation | Tooth, Alveolar bone | 50 |
| Rahman et al. (2024) | 2024 | Panoramic | Object Detection | Tooth, Caries, Infection, Impacted tooth, Fractured tooth, Broken down crown/root | 232 |
| Liu et al. (2023a) | 2023 | CBCT, 3D Intraoral | Semantic Segmentation | Tooth, Jaw | 503 pairs |
| Tufts Dental Database (Panetta et al. 2022) | 2022 | Panoramic | Semantic Segmentation | Tooth, maxillomandibular | 1,000 |
| NKUT (Zhou et al. 2024) | 2024 | CBCT | Semantic Segmentation | Wisdom tooth | 133 |
| ODSI-DB (Hyttinen et al. 2020) | 2020 | Spectral Oral Image | Semantic Segmentation | 35 classes including soft tissues, hard tissues, *etc*. | 316 |
| Román et al. (2021) | 2021 | Panoramic | - | - | 598 unlabeled |

Dataset. 2023. [accessed 2024 Nov]. https://github.com/Niihhaa/Dataset.

Ahn JS, Cho YR. 2024. Weighted sparse convolution and transformer feature aggregation networks for 3D dental segmentation. IEEE Access. 12:135172-135184.

Almalki A, Latecki LJ. 2024. Self-supervised learning with masked autoencoders for teeth segmentation from intra-oral 3D scans. Paper presented at: Proc IEEE Winter Conf Appl Comput Vis.

Tooth numbering computer vision project. 2024. [accessed 2024 Nov]. https://universe.roboflow.com/azleen-63bso/tooth-numbering.

Oral cancer (lips and tongue) images. 2020. [accessed 2024 Nov]. https://www.kaggle.com/datasets/shivam17299/oral-cancer-lips-and-tongue-images.

Ben-Hamadou A, Smaoui O, Chaabouni-Chouayakh H, Rekik A, Pujades S, Boyer E, Strippoli J, Thollot A, Setbon H, Trosset C. Forthcoming 2022. Teeth3DS: A benchmark for teeth segmentation and labeling from intra-oral 3d scans.

Ben-Hamadou A, Smaoui O, Rekik A, Pujades S, Boyer E, Lim H, Kim M, Lee M, Chung M, Shin Y-G et al. Forthcoming 2023 2023. 3DTeethSeg'22: 3D teeth scan segmentation and labeling challenge.

Brahmi W, Jdey I. 2024. Automatic tooth instance segmentation and identification from panoramic X-ray images using deep CNN. Multimedia Tools and Applications. 83(18):55565-55585.

Calazans MAA, Ferreira FABS, Alcoforado MdLMG, Santos Ad, Pontual AdA, Madeiro F. 2022. Automatic classification system for periapical lesions in cone-beam computed tomography. Sensors. 22(17):6481.

Cha J-Y, Yoon H-I, Yeo I-S, Huh K-H, Han J-S. 2021. Peri-implant bone loss measurement using a region-based convolutional neural network on dental periapical radiographs. J Clin Med. 10(5).

Chang H-J, Lee S-J, Yong T-H, Shin N-Y, Jang B-G, Kim J-E, Huh K-H, Lee S-S, Heo M-S, Choi S-C et al. 2020. Deep learning hybrid method to automatically diagnose periodontal bone loss and stage periodontitis. Scientific Rep. 10(1):7531.

Chen H, Qu Z, Tian Y, Jiang N, Qin Y, Gao J, Zhang R, Ma Y, Jin Z, Zhai G. 2024a. A cross-temporal multimodal fusion system based on deep learning for orthodontic monitoring. Comput Biol Med. 180:109025.

Chen H, Zhang K, Lyu P, Li H, Zhang L, Wu J, Lee C-H. 2019. A deep learning approach to automatic teeth detection and numbering based on object detection in dental periapical films. Scientific Rep. 9(1):3840.

Chen Q, Zhao Y, Liu Y, Sun Y, Yang C, Li P, Zhang L, Gao C. 2021. MSLPNet: Multi-scale location perception network for dental panoramic X-ray image segmentation. Neural Comput Appl. 33(16):10277-10291.

Chen R, Ma Y, Chen N, Liu L, Cui Z, Lin Y, Wang W. 2022. Structure-aware long short-term memory network for 3D cephalometric landmark detection. IEEE Trans Med Imag. 41(7):1791-1801.

Chen Y, Du H, Yun Z, Yang S, Dai Z, Zhong L, Feng Q, Yang W. 2020. Automatic segmentation of individual tooth in dental CBCT images from tooth surface map by a multi-task FCN. IEEE Access. 8:97296-97309.

Chen YC, Wang KH, Lin CL. 2024b. Personalized prosthesis design in all-on-4® treatment through deep learning-accelerated structural optimization. J Dent Sci. 19(4):2140-2149.

Choi J, Eun H, Kim C. 2018. Boosting proximal dental caries detection via combination of variational methods and convolutional neural network. J Signal Process Syst. 90(1):87-97.

Chung M, Lee J, Park S, Lee M, Lee CE, Lee J, Shin Y-G. 2021. Individual tooth detection and identification from dental panoramic X-ray images via point-wise localization and distance regularization. Artif Intell Med. 111:101996.

Chung M, Lee M, Hong J, Park S, Lee J, Lee J, Yang I-H, Lee J, Shin Y-G. 2020. Pose-aware instance segmentation framework from cone beam CT images for tooth segmentation. Comput Biol Med. 120:103720.

Cipriano M, Allegretti S, Bolelli F, Pollastri F, Grana C. 2022. Improving segmentation of the inferior alveolar nerve through deep label propagation. Paper presented at: Proc IEEE Conf Comput Vis Pattern Recog. New Orleans, LA, USA.

Costa ED, Gaêta-Araujo H, Carneiro JA, Zancan BAG, Baranauskas JA, Macedo AA, Tirapelli C. 2024. Development of a dental digital data set for research in artificial intelligence: The importance of labeling performed by radiologists. Oral Surg Oral Med Oral Pathol Oral Radiol. 138(1):205-213.

Cui W, Wang Y, Li Y, Song D, Zuo X, Wang J, Zhang Y, Zhou H, Chong BS, Zeng L et al. 2022a. CTooth+: A large-scale dental cone beam computed tomography dataset and benchmark for tooth volume segmentation. Paper presented at: Med Imag Comput Comput Assist Interv Workshop Data Augmentation Labeling Imperfections. Singapore.

Cui W, Wang Y, Zhang Q, Zhou H, Song D, Zuo X, Jia G, Zeng L. 2022b. CTooth: A fully annotated 3D dataset and benchmark for tooth volume segmentation on cone beam computed tomography images. Paper presented at: Int Conf Intell Robot Appl. Harbin, China.

Cui Z, Fang Y, Mei L, Zhang B, Yu B, Liu J, Jiang C, Sun Y, Ma L, Huang J et al. 2022c. A fully automatic ai system for tooth and alveolar bone segmentation from cone-beam ct images. Nature Communications. 13(1):2096.

Cui Z, Li C, Chen N, Wei G, Chen R, Zhou Y, Shen D, Wang W. 2021. TSegNet: An efficient and accurate tooth segmentation network on 3D dental model. Med Image Anal. 69:101949.

Cui Z, Li C, Wang W. 2019. ToothNet: Automatic tooth instance segmentation and identification from cone beam CT images. Paper presented at: Proc IEEE Conf Comput Vis Pattern Recog. Long Beach, CA, USA.

Danks RP, Bano S, Orishko A, Tan HJ, Moreno Sancho F, D'Aiuto F, Stoyanov D. 2021. Automating periodontal bone loss measurement via dental landmark localisation. Int J Comput Assisted Radiol Surgery. 16(7):1189-1199.

Ding W, Sun K, Yu M, Lin H, Feng Y, Li J, Liu Z. 2024. Accurate estimation of 6-DOF tooth pose in 3D intraoral scans for dental applications using deep learning. Frontiers Information Technol Electron Eng. 25(9):1240-1249.

Teethdecay. 2022. [accessed 2024 Mar]. https://www.kaggle.com/datasets/snginh/teethdecay.

Do HV, Vo TNN, Nguyen PT, Luong THL, Cu NG, Le HS. 2024. A dataset of apical periodontitis lesions in panoramic radiographs for deep-learning-based classification and detection. Data in Brief. 54:110486.

Du M, Liang K, Zhang L, Gao H, Liu Y, Xing Y. 2023. Deep-learning-based metal artefact reduction with unsupervised domain adaptation regularization for practical ct images. IEEE Trans Med Imag. 42(8):2133-2145.

Dental-caries-segmentation. 2022. [accessed 2024 Mar]. https://github.com/anumfatima427/Dental-Caries-Segmentation.

Gao S, Li X, Li X, Li Z, Deng Y. 2022. Transformer based tooth classification from cone-beam computed tomography for dental charting. Comput Biol Med. 148:105880.

Object_detection_on_cavities. 2022. [accessed 2024 Mar]. https://github.com/atul-g/object_detection_on_cavities.

Gou M, Rao Y, Zhang M, Sun J, Cheng K. 2019. Automatic image annotation and deep learning for tooth CT image segmentation. Paper presented at: Int Conf Image Graph. Beijing, China.

Haghanifar A, Majdabadi MM, Ko S-B. Forthcoming 2020 2020. PaXNet: Dental caries detection in panoramic X-ray using ensemble transfer learning and capsule classifier.

Hamamci IE, Er S, Simsar E, Yuksel AE, Gultekin S, Ozdemir SD, Yang K, Li HB, Pati S, Stadlinger B et al. Forthcoming 2023 2023. DENTEX: An abnormal tooth detection with dental enumeration and diagnosis benchmark for panoramic X-rays.

Hao J, Wong LM, Shan Z, Ai QYH, Shi X, Tsoi JKH, Hung KF. 2024. A semi-supervised transformer-based deep learning framework for automated tooth segmentation and identification on panoramic radiographs. Diagnostics (Basel). 14(17).

Hou S, Zhou T, Liu Y, Dang P, Lu H, Shi H. 2023. Teeth U-Net: A segmentation model of dental panoramic x-ray images for context semantics and contrast enhancement. Comput Biol Med. 152:106296.

Hsu T-MH, Wang Y-CC. 2021. DeepOPG: Improving orthopantomogram finding summarization with weak supervision. Paper presented at: Med Imag Comput Comput Assist Interv Proc. Strasbourg, France.

Huang Y, Liu W, Yao C, Miao X, Guan X, Lu X, Liang X, Ma L, Tang S, Zhang Z et al. 2024. A multimodal dental dataset facilitating machine learning research and clinic services. Sci Data. 11(1).

Hwang I-K, Kang S-R, Yang S, Kim J-M, Kim J-E, Huh K-H, Lee S-S, Heo M-S, Yi W-J, Kim T-I. 2023. SinusC-Net for automatic classification of surgical plans for maxillary sinus augmentation using a 3D distance-guided network. Scientific Rep. 13(1):11653.

Hwang J-J, Azernikov S, Efros A, Yu S. Forthcoming 2018 2018. Learning beyond human expertise with generative models for dental restorations.

Hyttinen J, Fält P, Jäsberg H, Kullaa A, Hauta-Kasari M. 2020. Oral and dental spectral image database—ODSI-DB. Appl Sci. 10(20):7246.

Jang TJ, Kim KC, Cho HC, Seo JK. 2022. A fully automated method for 3D individual tooth identification and segmentation in dental CBCT. IEEE Trans Pattern Anal Mach Intell. 44(10):6562-6568.

Jeoun B-S, Yang S, Lee S-J, Kim T-I, Kim J-M, Kim J-E, Huh K-H, Lee S-S, Heo M-S, Yi W-J. 2022. Canal-Net for automatic and robust 3D segmentation of mandibular canals in CBCT images using a continuity-aware contextual network. Scientific Rep. 12(1):13460.

Jiang H, Zhang P, Che C, Jin B. 2021. RDFNet: A fast caries detection method incorporating transformer mechanism. Comput Math Methods Med. 2021:9773917.

Jiang H, Zhang P, Che C, Jin B, Zhu Y. 2023. CariesFG: A fine-grained RGB image classification framework with attention mechanism for dental caries. Eng Appl Artif Intell. 123:106306.

Teeth detection and numbering computer vision project. 2023. [accessed 2024 Nov]. https://universe.roboflow.com/prime-snf1v/teeth-detection-and-numbering-agi2i.

Kalla M-P, Vagenas TP, Economopoulos TL, Matsopoulos GK. 2023. Deep learning-based registration of two-dimensional dental images with edge specific loss. J Med Imag. 10(3):034002.

Kim J, Lee H-S, Song I-S, Jung K-H. 2019. Dentnet: Deep neural transfer network for the detection of periodontal bone loss using panoramic dental radiographs. Scientific Rep. 9(1):17615.

Kong Z, Xiong F, Zhang C, Fu Z, Zhang M, Weng J, Fan M. 2020. Automated maxillofacial segmentation in panoramic dental X-ray images using an efficient encoder-decoder network. IEEE Access. 8:207822-207833.

Kunt L, Kybic J, Nagyová V, Tichý A. 2023. Automatic caries detection in bitewing radiographs: Part i--deep learning. Clin Oral Investigations. 27(12):7463-7471.

Calculus. 2023. [accessed 2024 Mar]. https://github.com/PKNU-PR-ML-Lab/calculus.

Lai Y, Fan F, Wu Q, Ke W, Liao P, Deng Z, Chen H, Zhang Y. 2021. LCANet: Learnable connected attention network for human identification using dental images. IEEE Trans Med Imag. 40(3):905-915.

Li D, Zhu M, Wang S, Hu Y, Yuan F, Yu J. 2024a. Accurate and automatic dental crown components segmentation with multi-scale attention based u-net and hybrid level set models. IEEE Trans Automat Sci Eng.1-12.

Li H, Zhou J, Zhou Y, Chen J, Gao F, Xu Y, Gao X. 2020a. Automatic and interpretable model for periodontitis diagnosis in panoramic radiographs. Paper presented at: Med Imag Comput Comput Assist Interv Proc. Lima, Peru.

Li K, Zhu J, Cui Z, Chen X, Liu Y, Wang F, Zhao Y. 2024b. A novel hierarchical cross-stream aggregation neural network for semantic segmentation of 3-D dental surface models. IEEE Trans Neural Netw Learn Syst. Pp.

Li P, Gao C, Liu F, Meng D, Yan Y. 2024c. THISNet: Tooth instance segmentation on 3D dental models via highlighting tooth regions. IEEE Trans Circuits Syst Video Technol. 34(7):5229-5241.

Li S, Pang Z, Song W, Guo Y, You W, Hao A, Qin H. 2020b. Low-shot learning of automatic dental plaque segmentation based on local-to-global feature fusion. Paper presented at: IEEE Int Symp Biomed Imag. Iowa City, IA, USA.

Li Y, Zeng G, Zhang Y, Wang J, Jin Q, Sun L, Zhang Q, Lian Q, Qian G, Xia N et al. 2022. AGMB-Transformer: Anatomy-guided multi-branch transformer network for automated evaluation of root canal therapy. IEEE J Biomed Health Inform. 26(4):1684-1695.

Lian C, Wang F, Deng HH, Wang L, Xiao D, Kuang T, Lin H-Y, Gateno J, Shen SGF, Yap P-T et al. 2020a. Multi-task dynamic transformer network for concurrent bone segmentation and large-scale landmark localization with dental CBCT. Paper presented at: Med Imag Comput Comput Assist Interv Proc. Lima, Peru.

Lian C, Wang L, Wu T-H, Wang F, Yap P-T, Ko C-C, Shen D. 2020b. Deep multi-scale mesh feature learning for automated labeling of raw dental surfaces from 3d intraoral scanners. IEEE Trans Med Imag. 39(7):2440-2450.

Lin Z, He Z, Wang X, Zhang B, Liu C, Su W, Tan J, Xie S. 2024. DBGANet: Dual-branch geometric attention network for accurate 3D tooth segmentation. IEEE Trans Circuits Syst Video Technol. 34(6):4285-4298.

Liu J, Hao J, Lin H, Pan W, Yang J, Feng Y, Wang G, Li J, Jin Z, Zhao Z et al. 2023a. Deep learning-enabled 3D multimodal fusion of cone-beam CT and intraoral mesh scans for clinically applicable tooth-bone reconstruction. Patterns. 4(9):100825.

Liu Y, Cheng Y, Song Y, Cai D, Zhang N. 2024a. Oral screening of dental calculus, gingivitis and dental caries through segmentation on intraoral photographic images using deep learning. BMC Oral Health. 24(1):1287.

Liu Y, Xia K, Cen Y, Ying S, Zhao Z. 2024b. Artificial intelligence for caries detection: A novel diagnostic tool using deep learning algorithms. Oral Radiol. 40(3):375-384.

Liu Z, He X, Wang H, Xiong H, Zhang Y, Wang G, Hao J, Feng Y, Zhu F, Hu H. 2023b. Hierarchical self-supervised learning for 3D tooth segmentation in intra-oral mesh scans. IEEE Trans Med Imag. 42(2):467-480.

Tooth segmentation on dental x-ray images. 2023. [accessed 2024 Nov]. https://www.kaggle.com/datasets/humansintheloop/teeth-segmentation-on-dental-x-ray-images.

Mahdi FP, Motoki K, Kobashi S. 2020. Optimization technique combined with deep learning method for teeth recognition in dental panoramic radiographs. Scientific Rep. 10(1):19261.

Dental radiography. 2023. [accessed 2024 Nov]. https://www.kaggle.com/datasets/imtkaggleteam/dental-radiography.

Nishitani Y, Nakayama R, Hayashi D, Hizukuri A, Murata K. 2021. Segmentation of teeth in panoramic dental X-ray images using U-Net with a loss function weighted on the tooth edge. Radiol Phys Technol. 14(1):64-69.

Ong SH, Kim H, Song JS, Shin TJ, Hyun HK, Jang KT, Kim YJ. 2024. Fully automated deep learning approach to dental development assessment in panoramic radiographs. BMC Oral Health. 24(1):426.

Panetta K, Rajendran R, Ramesh A, Rao SP, Agaian S. 2022. Tufts dental database: A multimodal panoramic x-ray dataset for benchmarking diagnostic systems. IEEE J Biomed Health Inform. 26(4):1650-1659.

Park S, Erkinov H, Hasan MAM, Nam S-H, Kim Y-R, Shin J, Chang W-D. 2023. Periodontal disease classification with color teeth images using convolutional neural networks. Electronics. 12(7).

Teeth_dataset. 2020. [accessed 2024 Mar]. https://www.kaggle.com/datasets/pushkar34/teeth-dataset.

Qian J, Cheng M, Tao Y, Lin J, Lin H. 2019. CephaNet: An improved Faster R-CNN for cephalometric landmark detection. Paper presented at: IEEE Int Symp Biomed Imag. Venice, Italy.

Qiu B, van der Wel H, Kraeima J, Glas HH, Guo J, Borra RJH, Witjes MJH, van Ooijen PMA. 2021. Mandible segmentation of dental CBCT scans affected by metal artifacts using coarse-to-fine learning model. J Personalized Med. 11(6).

Qiu L, Ye C, Chen P, Liu Y, Han X, Cui S. 2022. DArch: Dental arch prior-assisted 3D tooth instance segmentation with weak annotations. Paper presented at: Proc IEEE Conf Comput Vis Pattern Recog. New Orleans, LA, USA.

Rad AE, Rahim MSM, Rehman A, Saba T. 2016. Digital dental X-ray database for caries screening. 3D Res. 7(2):18.

Rahman RB, Tanim SA, Alfaz N, Shrestha TE, Miah MSU, Mridha MF. 2024. A comprehensive dental dataset of six classes for deep learning based object detection study. Data in Brief. 57:110970.

Rashid U, Javid A, Khan AR, Liu L, Ahmed A, Khalid O, Saleem K, Meraj S, Iqbal U, Nawaz R. 2022. A hybrid Mask RCNN-based tool to localize dental cavities from real-time mixed photographic images. PeerJ Comput Sci. 8:e888.

Raudaschl PF, Zaffino P, Sharp GC, Spadea MF, Chen A, Dawant BM, Albrecht T, Gass T, Langguth C, Lüthi M et al. 2017. Evaluation of segmentation methods on head and neck CT: Auto-segmentation challenge 2015. Med Phys. 44(5):2020-2036.

Román JCM, Fretes VR, Adorno CG, Silva RG, Noguera JLV, Legal-Ayala H, Mello-Román JD, Torres RDE, Facon J. 2021. Panoramic dental radiography image enhancement using multiscale mathematical morphology. Sensors. 21(9):3110.

Shen X, Zhang C, Jia X, Li D, Liu T, Tian S, Wei W, Sun Y, Liao W. 2023. TransDFNet: Transformer-based truncated signed distance fields for the shape design of removable partial denture clasps. IEEE J Biomed Health Inform. 27(10):4950-4960.

Shen Z, Shang X, Li Y, Bao Y, Zhang X, Dong X, Wan L, Xiong G, Wang F-Y. 2019a. PredNet and CompNet: Prediction and high-precision compensation of in-plane shape deformation for additive manufacturing. Paper presented at: IEEE Int Conf Automat Sci Eng. Vancouver, Canada.

Shen Z, Shang X, Zhao M, Dong X, Xiong G, Wang F-Y. 2019b. A learning-based framework for error compensation in 3D printing. IEEE Trans Cybern. 49(11):4042-4050.

Shi Y, Ye Z, Guo J, Tang Y, Dong W, Dai J, Miao Y, You M. 2024. Deep learning methods for fully automated dental age estimation on orthopantomograms. Clin Oral Investig. 28(3):198.

Silva G, Oliveira L, Pithon M. 2018. Automatic segmenting teeth in x-ray images: Trends, a novel data set, benchmarking and future perspectives. Expert Syst with Appl. 107:15-31.

Tan ZQ, Zhu X, Addison O, Li Y. 2025a. U-Mamba2: Scaling State Space Models for Dental Anatomy Segmentation in CBCT. Paper presented at: Medical Image Computing and Computer Assisted Intervention, Workshop on Oral and Dental Image Analysis. Daejeon, South Korea.

Tan ZQ, Zhu X, Addison O, Li Y. 2025b. U-Mamba2-SSL for Semi-Supervised Tooth and Pulp Segmentation in CBCT. Paper presented at: Medical Image Computing and Computer Assisted Intervention, Workshop on Oral and Dental Image Analysis. Daejeon, South Korea.

Thalji N, Aljarrah E, Almomani MH, Raza A, Migdady H, Abualigah L. 2024. Segmented X-ray image data for diagnosing dental periapical diseases using deep learning. Data in Brief. 54.

Tian S, Dai N, Zhang B, Yuan F, Yu Q, Cheng X. 2019. Automatic classification and segmentation of teeth on 3D dental model using hierarchical deep learning networks. IEEE Access. 7:84817-84828.

Tian S, Wang M, Dai N, Ma H, Li L, Fiorenza L, Sun Y, Li Y. 2022. DCPR-GAN: Dental crown prosthesis restoration using two-stage generative adversarial networks. IEEE J Biomed Health Inform. 26(1):151-160.

Tian S, Wang M, Yuan F, Dai N, Sun Y, Xie W, Qin J. 2021. Efficient computer-aided design of dental inlay restoration: A deep adversarial framework. IEEE Trans Med Imag. 40(9):2415-2427.

Torosdagli N, Anwar S, Verma P, Liberton DK, Lee JS, Han WW, Bagci U. 2023. Relational reasoning network for anatomical landmarking. J Med Imag. 10(2):24002.

Torosdagli N, Liberton DK, Verma P, Sincan M, Lee JS, Bagci U. 2019. Deep geodesic learning for segmentation and anatomical landmarking. IEEE Trans Med Imag. 38(4):919-931.

Vinayahalingam S, Goey R-S, Kempers S, Schoep J, Cherici T, Moin DA, Hanisch M. 2021. Automated chart filing on panoramic radiographs using deep learning. J Dentistry. 115:103864.

Cephalometric landmark detection in lateral x-ray images 2023. 2023a. [accessed 2024 Nov]. https://cl-detection2023.grand-challenge.org/.

Wang C-W, Huang C-T, Lee J-H, Li C-H, Chang S-W, Siao M-J, Lai T-M, Ibragimov B, Vrtovec T, Ronneberger O et al. 2016. A benchmark for comparison of dental radiography analysis algorithms. Med Image Anal. 31:63-76.

Wang X, Gao S, Jiang K, Zhang H, Wang L, Chen F, Yu J, Yang F. 2023b. Multi-level uncertainty aware learning for semi-supervised dental panoramic caries segmentation. Neurocomputing. 540:126208.

Wirtz A, Mirashi SG, Wesarg S. 2018. Automatic teeth segmentation in panoramic X-ray images using a coupled shape model in combination with a neural network. Paper presented at: Med Imag Comput Comput Assist Interv Proc. Granada, Spain.

Wu T-H, Lian C, Lee S, Pastewait M, Piers C, Liu J, Wang F, Wang L, Chiu C-Y, Wang W et al. 2022. Two-stage mesh deep learning for automated tooth segmentation and landmark localization on 3D intraoral scans. IEEE Trans Med Imag. 41(11):3158-3166.

Xie R, Yang Y, Chen Z. 2023. WITS: Weakly-supervised individual tooth segmentation model trained on box-level labels. Pattern Recognit. 133:108974.

Xu X, Liu C, Zheng Y. 2019. 3D tooth segmentation and labeling using deep convolutional neural networks. IEEE Trans Vis Comput Graph. 25(7):2336-2348.

Xu Z, Wan P, Aihemaiti G, Zhang D. 2021. Exploiting saliency in attention based convolutional neural network for classification of vertical root fractures. Paper presented at: Proc Int Conf Pattern Recognit Workshop.

Xue T, Chen L, Sun Q. 2024. Deep learning method to automatically diagnose periodontal bone loss and periodontitis stage in dental panoramic radiograph. J Dent. 150:105373.

Yang X, Xie J, Li X, Li X, Shen L, Deng Y. 2023. TCSIoT: Text guided 3d context and slope aware triple network for dental implant position prediction. Paper presented at: IEEE Int Conf Bioinf Biomed. Instanbul, Turkey.

Ying S, Wang B, Zhu H, Liu W, Huang F. 2022. Caries segmentation on tooth X-ray images with a deep network. J Dentistry. 119:104076.

Zanjani FG, Anssari Moin D, Verheij B, Claessen F, Cherici T, Tan T, de With PHN. 2019. Deep learning approach to semantic segmentation in 3D point cloud intra-oral scans of teeth. Paper presented at: Proc Int Conf Med Imag Deep Learn. London, United Kingdom.

Zanjani FG, Pourtaherian A, Zinger S, Moin DA, Claessen F, Cherici T, Parinussa S, de With PHN. 2021. Mask-MCNet: Tooth instance segmentation in 3D point clouds of intra-oral scans. Neurocomputing. 453:286-298.

Zeng M, Yan Z, Liu S, Zhou Y, Qiu L. 2021. Cascaded convolutional networks for automatic cephalometric landmark detection. Med Image Anal. 68:101904.

Zhang F, Zhu J, Hao P, Wu F, Zheng Y. 2022a. BDU-Net: Toward accurate segmentation of dental image using border guidance and feature map distortion. Int J Imag Syst Technol. 32(4):1221-1230.

Zhang J, Liu M, Wang L, Chen S, Yuan P, Li J, Shen SG-F, Tang Z, Chen K-C, Xia JJ et al. 2017. Joint craniomaxillofacial bone segmentation and landmark digitization by context-guided fully convolutional networks. Paper presented at: Med Imag Comput Comput Assist Interv Proc. Quebec, Canada.

Zhang K, Chen H, Lyu P, Wu J. 2022b. A relation-based framework for effective teeth recognition on dental periapical X-rays. Computerized Med Imag Graph. 95:102022.

Zhang K, Wu J, Chen H, Lyu P. 2018. An effective teeth recognition method using label tree with cascade network structure. Computerized Med Imag Graph. 68:61-70.

Zhang M, Zhao N, Zhuang Y, Wang L, Tao X. 2024. FaLdViT: A simple yet effective framework to detect cephalometric landmarks. Paper presented at: Proc Int Conf Comput Supported Cooperative Work Des.

Zhang Y, Ye F, Chen L, Xu F, Chen X, Wu H, Cao M, Li Y, Wang Y, Huang X. 2023. Children's dental panoramic radiographs dataset for caries segmentation and dental disease detection. Scientific Data. 10(1):380.

Zhao M, Xiong G, Fang Q, Dong X, Wang F, Han Y, Shen Z, Wang FY. 2024. Enlarge the error prediction dataset in 3-D printing: An unsupervised dental crown mesh generator. IEEE Trans Comput Social Syst. 11(6):7929-7940.

Zhao Y, Li P, Gao C, Liu Y, Chen Q, Yang F, Meng D. 2020. TSASNet: Tooth segmentation on dental panoramic X-ray images by two-stage attention segmentation network. Knowledge-Based Syst. 206:106338.

Zhao Y, Zhang L, Liu Y, Meng D, Cui Z, Gao C, Gao X, Lian C, Shen D. 2022. Two-stream graph convolutional network for intra-oral scanner image segmentation. IEEE Trans Med Imag. 41(4):826-835.

Zheng Y, Chen B, Shen Y, Shen K. 2023. TeethGNN: Semantic 3D teeth segmentation with graph neural networks. IEEE Trans Vis Comput Graph. 29(7):3158-3168.

Zheng Z, Yan H, Setzer FC, Shi KJ, Mupparapu M, Li J. 2021. Anatomically constrained deep learning for automating dental CBCT segmentation and lesion detection. IEEE Trans Automat Sci Eng. 18(2):603-614.

Zhou X, Yu G, Yin Q, Yang J, Sun J, Lv S, Shi Q. 2023. Tooth type enhanced transformer for children caries diagnosis on dental panoramic radiographs. Diagnostics. 13(4).

Zhou Z, Chen Y, He A, Que X, Wang K, Yao R, Li T. 2024. NKUT: Dataset and benchmark for pediatric mandibular wisdom teeth segmentation. IEEE J Biomed Health Inform. 28(6):3523-3533.

Zhu H, Cao Z, Lian L, Ye G, Gao H, Wu J. 2023. Cariesnet: A deep learning approach for segmentation of multi-stage caries lesion from oral panoramic X-ray image. Neural Comput Appl. 35(22):16051-16059.
